# OCTAL (Oxford Cognitive Testing Portal): A remote, cross-cultural cognitive assessment detects domain-specific aging and dementia

**DOI:** 10.1101/2025.06.23.25330153

**Authors:** Sijia Zhao, Sofia Toniolo, Qian-Yuan Tang, Anna Scholcz, Akke Ganse-Dumrath, Claudia Gendarini, M. John Broulidakis, Sian Thompson, Sanjay G. Manohar, Masud Husain

**Author notes:** Corresponding Authors:* Sijia Zhao, Department of Experimental Psychology, New Radcliffe House, Walton Street, Oxford OX2 6GG, United Kingdom. SZ and ST had equal contribution to this paper.

## Abstract

The global rise in dementia necessitates scalable cognitive assessments that can evolve to serve both clinical and research applications. We present the Oxford Cognitive Testing Portal (OCTAL), a remote, browser-based platform providing performance metrics for memory, attention, visuospatial and executive function domains. Four validation studies (N=1,749) confirmed cross-cultural applicability, lifespan sensitivity and clinical utility. Task performance was equivalent in English- and Chinese-speaking younger adults and mapped domain-specific ageing trajectories in mid- to late-adulthood. In a memory-clinic cohort (N=194), a 5-minute OCTAL screen distinguished patients with Alzheimer’s disease dementia from subjective cognitive decline (AUC = 0.92), matching a standard paper-based test, while a 20-minute subset surpassed this (AUC = 0.98; p = 0.04). Test-retest reliability was very good (ICC ≥ 0.79; N = 118). OCTAL enables remote assessment for large-scale research and screening, with an open, modular architecture that makes it a uniquely sustainable and evolvable tool for the research community.

## Introduction

Dementia prevalence is increasing worldwide as global populations age, while capacity of memory assessment services has not kept pace.^1–3^ In the United States alone, 7.2 million adults aged ≥ 65 are currently living with Alzheimer’s disease (AD), a figure projected to almost double by 2060.^4^ Meanwhile, workforce shortages in primary-care and specialist memory clinics, coupled with linguistic and cultural barriers, deprive many individuals of timely cognitive assessment and longitudinal monitoring.^3^ There is also increasing recognition that screening for AD and other neurodegenerative conditions at scale might be important to introduce interventions – pharmacological and lifestyle – that might delay progression to dementia onset.^5^

Public demand for proactive cognitive evaluation is equally compelling, as reflected in a recent survey of 1,702 US adults over 45, who were demographically matched to U.S. households^4^. The results showed that 99 % view an early diagnosis of AD as important, while 59% believe routine cognitive screening is a vital part of preventive care.^4^ These attitudes create both an ethical imperative and a market pull for tests that (i) detect impairment with gold-standard accuracy, (ii) can be administered repeatedly without ceiling or practice effects, and (iii) scale via unsupervised, browser-based delivery acceptable to culturally diverse users.

Yet, the current gold-standard cognitive screening tests — the Mini-Mental State Examination (MMSE),^6^ Montreal Cognitive Assessment (MoCA)^7^ and Addenbrooke’s Cognitive Examination-III (ACE-III)^8^ — remain almost exclusively pen-and-paper. Each administration occupies 15–20 minutes of clinician time, limiting patient throughput in already overstretched services and, crucially, lengthening patients’ waiting times.^9^ Despite their clinical utility, their coarse scoring offers limited granularity for tracking subtle decline, particularly in individuals with high baseline functioning.^10,11^ Further, the format is poorly suited to population-level screening and longitudinal monitoring; the requirement for in-person administration prevents seamless integration with emerging fluid biomarker pipelines, which are increasingly designed for scalable, remote deployment (e.g., AD-related plasma biomarker assays).^12^

Remote, browser-based testing can circumvent these constraints while enriching psychometrics.^9,13,14^ Digital platforms capture millisecond-level reaction-time precision and log every interaction during task execution, providing a complete behavioural trajectory that enables fine-grained cognitive stratification and supports the high-frequency sampling required to detect within-person change.^10^

Here, we introduce the Oxford Cognitive Testing Portal (OCTAL), a fully remote, browser-based cognitive assessment platform optimised for personal devices ^1^ (Figure 1). OCTAL comprises scientifically grounded, user-friendly tasks measuring memory, attention, executive function, and visuospatial processing; optional secondary modules can be appended to address specific research questions.^15,16^

**Figure 1:**
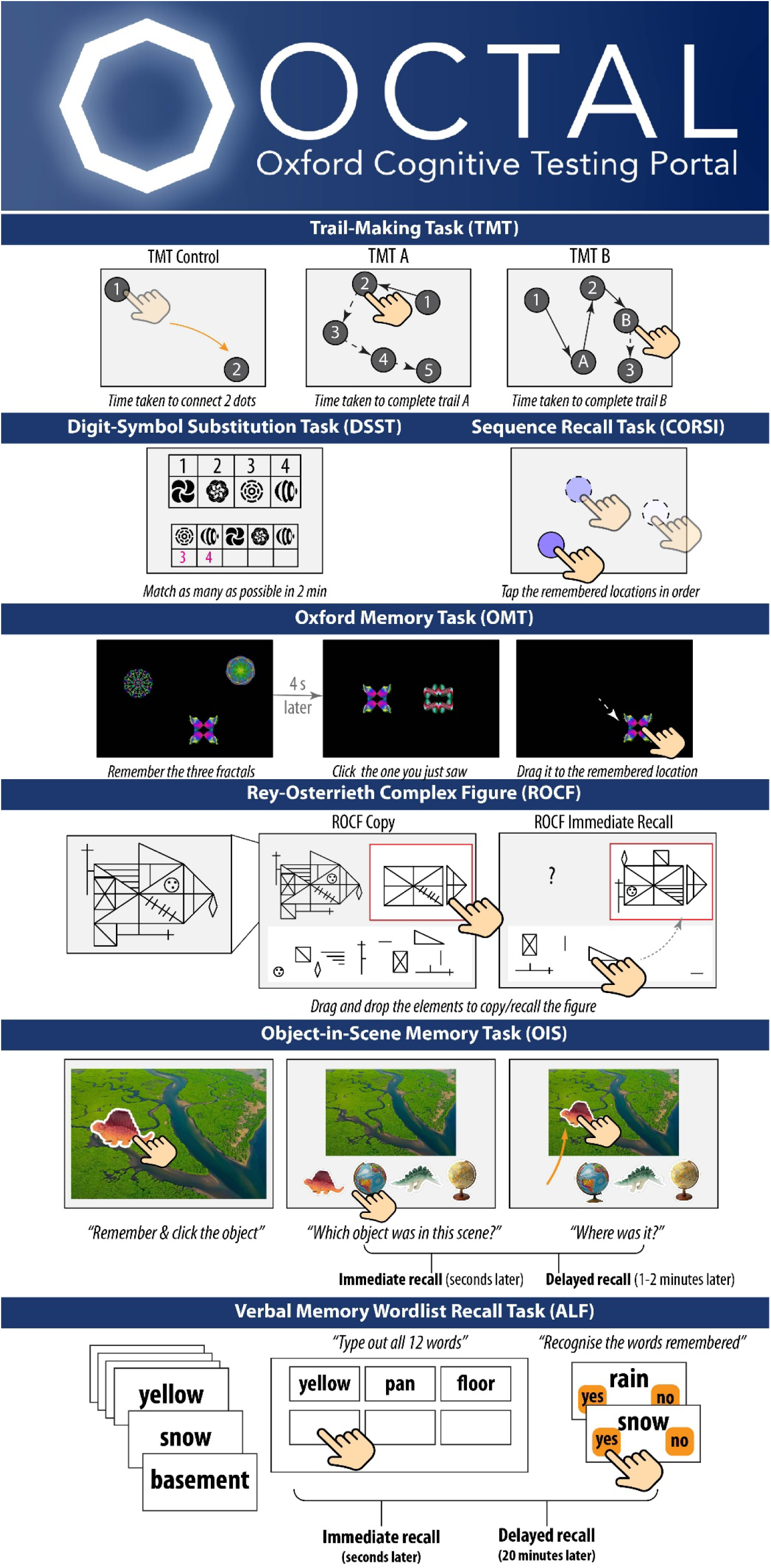
OCTAL (Oxford Cognitive Testing Portal) task battery and outcome measures. 1) **Trail-Making Task (TMT)** — Participants tap onscreen nodes as rapidly as possible. A two-dot control (‘Connect-2-Dots’) gauges basic psychomotor speed. They then complete two 25-node numeric trails (TMT-A, a measure of attention and processing speed) and two alternating number-letter trails (TMT-B, a measure of the ability to switch between stimuli categories). Metrics are mean completion time for each condition (TMT_Connect2Dots, TMT_A, TMT_B) and the TMT_B/A ratio, a marker of executive function (cognitive flexibility). 2) **Digit-Symbol Substitution Task (DSST)** — Guided by a reference key, participants match as many symbols with correct digits as possible in 2 min. Outcomes are mean reaction time for correct substitutions (DSST_RT) and proportion correct (DSST_Accuracy). This tests attention and processing speed. 3) **Sequence Recall Task (CORSI)** — After observing a sequence of one, two or three highlighted circles, participants reproduce the order. Performance is summarised by mean localisation error (CORSI_LocErr) and between-trial coefficient of variation (CORSI_CoV). This tests spatial short-term memory. 4) **Oxford Memory Task (OMT)** — Participants view either one (easy) or three (hard) fractals and, after 4 s, drag the target fractal to its original location. Metrics are recognition accuracy (OMT_Accuracy) and spatial displacement error (OMT_LocError). This tests object recognition and spatial short-term memory. 5) **Rey–Osterrieth Complex Figure (ROCF)** — Participants drag-and-drop 13 predefined elements to recreate a figure (“copy”) and then reproduce it from memory immediately afterwards (“recall”). Element placement is evaluated with pixel-level precision relative to a fixed central anchor, yielding proportional scores for copy (ROCF_CopyScore which is a measure of visuospatial perception and visuoconstructive skills) and recall (ROCF_RecallScore which is a measure of visuospatial episodic memory) on a continuous 0–100% scale; this extends the conventional 0–36 discrete system by incorporating spatial accuracy. Memory retention, adjusted for individual visuomotor or perceptual demands, is expressed as the ratio ROCF_Remember = ROCF_RecallScore ⁄ ROCF_CopyScore in percentage. See Methods, ‘Ethical and intellectual property considerations’, for stimulus rationale and copyright details 6) **Object-in-Scene Memory Task (OIS**) —participants identify an object placed on a random location on a natural scene and subsequently choose the object and report its location after a short (seconds) and longer (1–2 min) delay. Metrics are recognition accuracy and localisation error for short-term memory (OIS_STM_Accuracy, OIS_STM_LocErr) and long-term memory (OIS_LTM_Accuracy, OIS_LTM_LocErr). 7) **Verbal Memory Wordlist Recall Task (ALF,** aka Accelerated Long-Term Forgetting) — After seeing a sequence of 12 words, participants freely recall items (ALF_nWords_immediate) and complete a yes/no recognition test (ALF_RecAccuracy_immediate); the procedure repeats 20 min later after intervening tasks (ALF_nWords_delayed, ALF_RecAcc_delayed). This is a measure of verbal episodic memory.

We report four validation studies (see Table 1 for demographic details of all four studies)

1. **Cross-cultural generalisability**. Deployment in English and Chinese among 363 young adults (18–45 years) in the United Kingdom and China demonstrated largely comparable performance.
2. **Lifespan scalability**. Data from 905 UK participants aged 45–85 years with cognitive domain-specific ageing trajectories.
3. **Clinical utility**. In a clinical cohort (AD, mild cognitive impairment [MCI], subjective cognitive decline [SCD]; N=194), OCTAL detected cognitive impairment defined by the ACE-III cut-off with high accuracy (AUC ≥ 0.90) and out-performed ACE-III in distinguishing AD/MCI from SCD.
4. **Test–retest reliability**. Re-administration up to three times over six months (N=118) showed good to excellent test-retest reliability that is comparable to ACE-III.

**Table 1:**
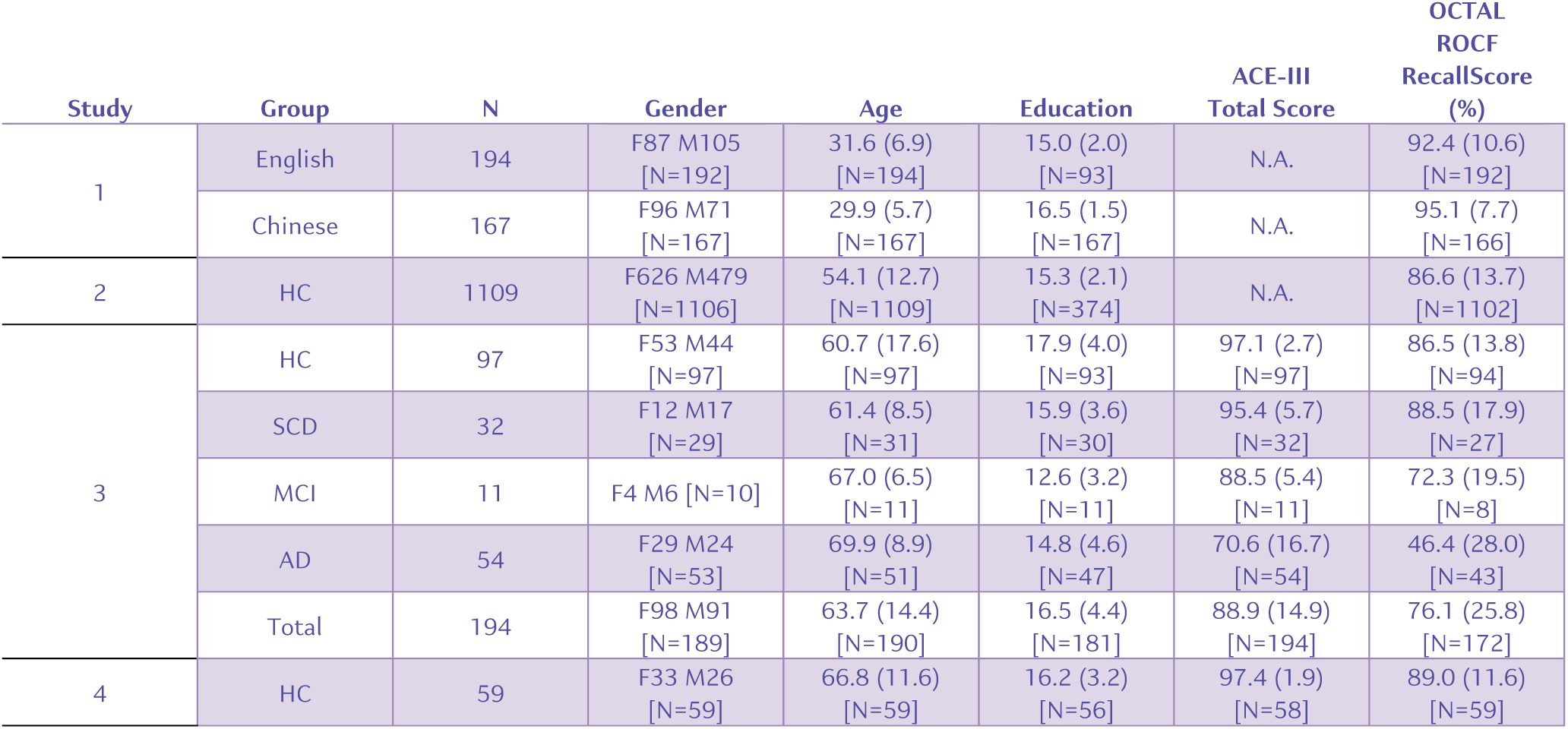
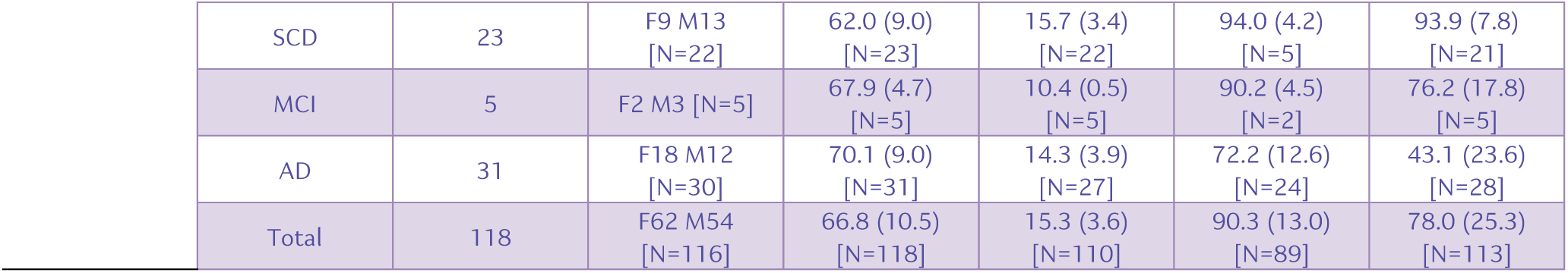
Demographic characteristics and ACE-III and OCTAL performance across the four studies. For the continuous variables (age, number of years education etc) data are presented as mean (1 standard deviation). Square brackets denote the number of participants with valid data (i.e., excluding the missing data). N.A. indicates data not applicable. For Study 4, ACE-III and OCTAL scores are averaged across all assessments. Further details are provided in the manuscript and Methods.

| Study | Group | N | Gender | Age | Education | ACE-III<br>Total Score | OCTAL<br>ROCF<br>RecallScore<br>(%) |
| --- | --- | --- | --- | --- | --- | --- | --- |
| 1 | English | 194 | F87 M105<br>[N=192] | 31.6 (6.9)<br>[N=194] | 15.0 (2.0)<br>[N=93] | N.A. | 92.4 (10.6)<br>[N=192] |
|  | Chinese | 167 | F96 M71<br>[N=167] | 29.9 (5.7)<br>[N=167] | 16.5 (1.5)<br>[N=167] | N.A. | 95.1 (7.7)<br>[N=166] |
| 2 | HC | 1109 | F626 M479<br>[N=1106] | 54.1 (12.7)<br>[N=1109] | 15.3 (2.1)<br>[N=374] | N.A. | 86.6 (13.7)<br>[N=1102] |
| 3 | HC | 97 | F53 M44<br>[N=97] | 60.7 (17.6)<br>[N=97] | 17.9 (4.0)<br>[N=93] | 97.1 (2.7)<br>[N=97] | 86.5 (13.8)<br>[N=94] |
|  | SCD | 32 | F12 M17<br>[N=29] | 61.4 (8.5)<br>[N=31] | 15.9 (3.6)<br>[N=30] | 95.4 (5.7)<br>[N=32] | 88.5 (17.9)<br>[N=27] |
|  | MCI | 11 | F4 M6 [N=10] | 67.0 (6.5)<br>[N=11] | 12.6 (3.2)<br>[N=11] | 88.5 (5.4)<br>[N=11] | 72.3 (19.5)<br>[N=8] |
|  | AD | 54 | F29 M24<br>[N=53] | 69.9 (8.9)<br>[N=51] | 14.8 (4.6)<br>[N=47] | 70.6 (16.7)<br>[N=54] | 46.4 (28.0)<br>[N=43] |
|  | Total | 194 | F98 M91<br>[N=189] | 63.7 (14.4)<br>[N=190] | 16.5 (4.4)<br>[N=181] | 88.9 (14.9)<br>[N=194] | 76.1 (25.8)<br>[N=172] |
| 4 | HC | 59 | F33 M26<br>[N=59] | 66.8 (11.6)<br>[N=59] | 16.2 (3.2)<br>[N=56] | 97.4 (1.9)<br>[N=58] | 89.0 (11.6)<br>[N=59] |

|  |  |  |  |  |  |  |
| --- | --- | --- | --- | --- | --- | --- |
| SCD | 23 | F9 M13<br>[N=22] | 62.0 (9.0)<br>[N=23] | 15.7 (3.4)<br>[N=22] | 94.0 (4.2)<br>[N=5] | 93.9 (7.8)<br>[N=21] |
| MCI | 5 | F2 M3 [N=5] | 67.9 (4.7)<br>[N=5] | 10.4 (0.5)<br>[N=5] | 90.2 (4.5)<br>[N=2] | 76.2 (17.8)<br>[N=5] |
| AD | 31 | F18 M12<br>[N=30] | 70.1 (9.0)<br>[N=31] | 14.3 (3.9)<br>[N=27] | 72.2 (12.6)<br>[N=24] | 43.1 (23.6)<br>[N=28] |
| Total | 118 | F62 M54<br>[N=116] | 66.8 (10.5)<br>[N=118] | 15.3 (3.6)<br>[N=110] | 90.3 (13.0)<br>[N=89] | 78.0 (25.3)<br>[N=113] |

Together, these findings demonstrate that OCTAL offers a practical, sensitive, and robust tool for digital screening and longitudinal monitoring, suitable for both clinical and research applications. Importantly, OCTAL is designed with researchers in mind. It produces data-rich outputs that are readily compatible with a wide range of digital platforms, including survey tools (e.g., Qualtrics), online participant recruitment systems (e.g., Prolific, MTurk), experimental testing platforms (e.g., Pavlovia, Gorilla), and complementary cognitive tools (e.g., Sea Hero Quest^17^, Cognitron^18^). OCTAL supports both remote and in-person deployment and enables high-resolution cognitive phenotyping across diverse populations and study designs.

## Study 1: International scalability in young adults

To first assess the feasibility and international scalability of OCTAL, and determine whether its tasks are inherently biased towards English speakers, we conducted the first study with 363 young adults below 45 years old (aged 18–44), including 194 native English speakers on an English version and 167 native Chinese speakers on a Chinese version of OCTAL^2^. All participants completed OCTAL remotely on their own laptops or desktop computers, regardless of operating system or browser. The platform was developed in PsychoJS, facilitating straightforward modification by researchers with minimal coding experience, and hosted on Pavlovia. Recruitment was conducted via Prolific (English version with UK residents) and Credamo (Chinese version with residents of mainland China). **Table 2** summarises demographic characteristics; no meaningful differences emerged between the two cohorts (except Chinese participants were slightly more educated).

**Table 2.** Demographic and cognitive performance comparisons between English- and Chinese-speaking participants (Study 1). Bootstrap-based practical significance was evaluated with a permutation procedure (10,000 iterations). For each metric, lower and upper thresholds were defined as 1 SD below and 1 SD above the mean of the English-speaking cohort, respectively. To determine whether the Chinese group (N = 167) differed meaningfully from these thresholds, N observations were resampled with replacement from the Chinese data, the mean was calculated, and this process was repeated 10,000 times. A difference was deemed significant when ≥97.5 % of resampled means lay below the lower threshold or above the upper threshold (p < 0.025). Non-parametric group comparison for sample mean (Mann–Whitney U tests) are reported in Supplementary Table 1. Reported values include the U statistics, the effect size (r) and Bonferroni-corrected p values.

| Metric | Mean (SD) | | Bootstrap $p$-value | |
| --- | --- | --- | --- | --- |
|  | English | Chinese | Below - 1SD | Above 1SD |
| $N$ | 194 | 167 | | |
| Gender (% Female) | 44.9 | 57.5 |  |  |
| Age (years) | 31.60 (6.88) | 29.88 (5.72) | N.S. | N.S. |
| Education (years) | 14.96 (2.02) | 16.46 (1.52) | N.S. | N.S. |
| ROCF_CopyScore | 99.07 (4.75) | 99.26 (2.36) | N.S. | N.S. |
| ROCF_CopyDuration | 105.87 (38.54) | 117.85 (52.62) | N.S. | N.S. |
| ROCF_RecallScore | 92.36 (10.60) | 95.07 (7.73) | N.S. | N.S. |
| ROCF_Remember | 93.15 (9.38) | 95.77 (7.36) | N.S. | N.S. |
| ROCF_RecallDuration | 88.92 (39.91) | 100.97 (56.53) | N.S. | N.S. |
| OMT_Acc_Easy | 96.95 (5.85) | 98.86 (2.46) | N.S. | N.S. |
| OMT_Acc_Hard | 85.88 (10.16) | 89.96 (8.76) | N.S. | N.S. |
| OMT_LocErr_Easy | 0.08 (0.05) | 0.07 (0.03) | N.S. | N.S. |
| OMT_LocErr_Hard | 0.16 (0.07) | 0.15 (0.07) | N.S. | N.S. |
| OMT_Acc | 91.42 (6.41) | 94.41 (4.84) | N.S. | N.S. |
| OMT_IdeRT | 1.73 (0.49) | 2.28 (0.79) | N.S. | N.S. |
| OMT_LocErr | 0.12 (0.05) | 0.11 (0.04) | N.S. | N.S. |
| OMT_LocRT | 2.10 (0.77) | 2.50 (1.86) | N.S. | N.S. |
| OMT_TargetDetection | 72.74 (13.84) | 82.12 (12.55) | N.S. | N.S. |
| OMT_Misbinding | 24.90 (10.91) | 17.92 (11.88) | N.S. | N.S. |
| OMT_Guessing | 14.73 (8.98) | 8.88 (7.16) | N.S. | N.S. |
| OIS_STM_SemanticAcc | 99.65 (2.86) | 99.88 (0.77) | N.S. | N.S. |
| *OIS_STM_Acc* | 97.33 (4.76) | 97.49 (7.68) | N.S. | N.S. |
| *OIS_STM_IdeRT* | 1.73 (0.24) | 2.57 (0.92) | N.S. | $p < 0.001$ |
| *OIS_STM_LocErr* | 0.21 (0.05) | 0.18 (0.05) | N.S. | N.S. |
| *OIS_STM_LocRT* | 5.71 (0.39) | 6.15 (3.64) | N.S. | N.S. |
| *OIS_LTM_SemanticAcc* | 86.11 (16.75) | 87.10 (15.28) | N.S. | N.S. |
| *OIS_LTM_Acc* | 77.94 (19.79) | 81.29 (18.42) | N.S. | N.S. |
| *OIS_LTM_IdeRT* | 6.65 (1.90) | 6.27 (1.82) | N.S. | N.S. |
| *OIS_LTM_LocErr* | 0.21 (0.05) | 0.20 (0.06) | N.S. | N.S. |
| *OIS_LTM_LocRT* | 11.18 (1.12) | 10.33 (3.49) | N.S. | N.S. |
| *DSST_nCorrectResponse* | 52.11 (14.08) | 51.88 (11.02) | N.S. | N.S. |
| *DSST_RT* | 2.37 (2.27) | 2.20 (0.47) | N.S. | N.S. |
| *DSST_pHit* | 97.91 (3.57) | 98.95 (1.90) | N.S. | N.S. |
| *TMT_Connect2* | 3.72 (0.98) | 2.56 (0.56) | N.S. | N.S. |
| *TMT_A* | 20.28 (9.24) | 20.78 (7.10) | N.S. | N.S. |
| *TMT_B* | 29.01 (11.54) | 31.53 (9.09) | N.S. | N.S. |
| *TMT_BdA* | 147.65 (35.30) | 156.64 (40.80) | N.S. | N.S. |

Cognitive performance on core OCTAL metrics was largely comparable between the English-speaking and Chinese-speaking groups (**Table 2**). Indeed, for the majority of key metrics, the mean performance of the Chinese cohort fell within the normative range established by the English-speaking cohort (defined as ±1 standard deviation of the English group’s mean, determined via a bootstrap permutation procedure; see **Table 2**).

However, when examining reaction times, a significant group difference that exceeded this normative threshold was observed for the identification response time in the immediate recall phase of the Object-in-Scene (OIS) task (OIS_STM_IdeRT). Chinese participants took an average of 2.57 seconds (SD=0.92) for this measure, compared to 1.73 seconds (SD=0.24) for English-speaking participants. This difference meant the Chinese group’s mean reaction time was significantly greater than one standard deviation above the English group’s mean. A speed-accuracy trade-off was an unlikely explanation for this specific finding, as the Chinese group showed only a marginal 0.16% advantage in OIS identification accuracy (97.49% vs. 97.33%), a difference too small to account for the substantial variance in response time. Meanwhile, although insignificant, several reaction time measures indicated slower responses in the Chinese group, the reason behind this systematically slower response is not immediately clear. We will come back to this in ***Discussion***.

A noteworthy difference emerged in Trail Making Test Part B (TMT-B), where Chinese participants were slower than the UK cohort (China: M = 31.5 s, SD = 9.1; UK: M = 29.0 s, SD = 11.4; U = 29,565, p = 0.01, r = 0.20), despite having learned English since childhood. Conversely, TMT-A performance was comparable between the groups (U = 31,307, p = 1.00, r = 0.10). The slower TMT-B performance in the Chinese cohort is likely attributable to the cognitive demands of switching between numerals and the English alphabet, a task inherently more demanding for second-language users. This hypothesis was strengthened by data from a second study involving 1,109 UK residents. In this sample, non-native English speakers (n=191) demonstrated significantly greater TMT-B latency from age 55 onwards relative to native speakers (n=918), an effect that persisted after controlling for TMT-A performance (see **Supplementary Materials**, “Does speaking English as a second language affect OCTAL performance?” and **Supplementary Figure 1**).

### Could the variance in OCTAL performance be merely explained by difference in devices?

During development of the platform we considered whether display size might confound OCTAL scores. The Rey–Osterrieth Complex Figure (ROCF; see Methods, ‘Ethical and intellectual property considerations’, for stimulus rationale and copyright details) was the prime candidate for such an effect: on a large screen, repeatedly dragging elements could prolong completion time, whereas on a small screen limited motor precision might hamper accurate placement. To isolate memory from such visuomotor factors we required participants to copy the figure first and then reproduce it from memory. Performance is therefore indexed both by the raw Copy score (i.e., a 100% scale showing how precise are ROCF’s elements located relative to each other) and by the Recall ⁄ Copy ratio, which normalises for any screen-related copying differences. To test this hypothesis, we expanded the mainland-Chinese cohort with 43 additional adults aged 45–70, yielding *N* = 210. All participants used either a laptop or a desktop computer; tablets were disallowed. Although device type and pointing method were not logged explicitly, the browser captured screen width and height (pixels) at test.

**Supplementary Figure 2** ranks the correlations between screen width and all 34 OCTAL metrics. Only ROCF Copy was affected: wider screens were associated with longer completion times (ρ = –0.26, *p* = 0.01) and higher precision (ρ = 0.26, *p* = 0.01). Time and accuracy were, however, independent (ρ = – 0.04, *p* = 0.61), discounting a simple speed–accuracy trade-off. **Supplementary Figure 3** shows analogous results for screen height. Copy score again correlated with screen height (ρ = 0.30, *p* = 0.0004), and the strength of the width and height associations did not differ (Fisher’s z = –1.60, *p* = 0.11). Time taken to complete ROCF copy, by contrast, was unrelated to height.

Crucially, how well ROCF was recalled (ROCF Recall score) did not correlate with screen dimensions (width ρ = 0.13; height ρ = 0.11; both *p* = 1). The memory index (Recall ⁄ Copy × 100 %) was likewise insensitive to screen size (width ρ = 0.06; height ρ = 0.02; both *p* = 1). Thus, while larger displays may marginally facilitate more precise copying—every participant nevertheless scored above 90 %—they do not influence the amount of information retained. OCTAL memory measures are therefore robust to the variability in laptop and desktop screen dimensions encountered in remote testing.

## Study 2: Feasibility of unsupervised testing and sensitivity to cognitive aging

Study 1 demonstrated that culturally diverse young adults can complete OCTAL unaided, demonstrating the platform’s scalability and accessibility amongst people below 45 years old in both UK and China. We next asked whether adults in mid- and late-life adults do the same and whether OCTAL detects changes of healthy ageing across cognitive domains. We recruited 1,109 UK residents via Prolific, of whom 905 were aged 45–89 years. All assessments were completed remotely without supervision, and only a few participants failed the attention checks (see Methods), indicating strong engagement and accessibility across the lifespan.

Analysis of cognitive performance revealed that while most metrics correlated significantly with age (**Supplementary Table 4**), the relationship was strongest for executive functions. The year-by-year trajectories (**Figure 2**) for executive tasks showed a continuous decline. On the Digit Symbol Substitution Test (DSST), reaction time increased steadily, eventually surpassing the boundary for abnormal performance (**Figure 2A-B**). Performance began to diverge from the young adult norm by age 46, but did not fall outside the normative range until age 56 (**Figure 2B**), with error rates remaining low until after age 65 (**Supplementary Figure 5A-B**). Similarly, completion times on the Trail Making Test (TMT-A and TMT-B) slowed progressively, breaching the abnormality threshold in the mid-to-late fifties (**Supplementary Figure 5E-H**). This executive decline was not an artefact of motor slowing or technological unfamiliarity, as performance on a visuomotor control task showed only a modest latency increase (**Supplementary Figure 5C**) and remained within normal limits across the lifespan (**Supplementary Figure 5D**).

**Figure 2:**
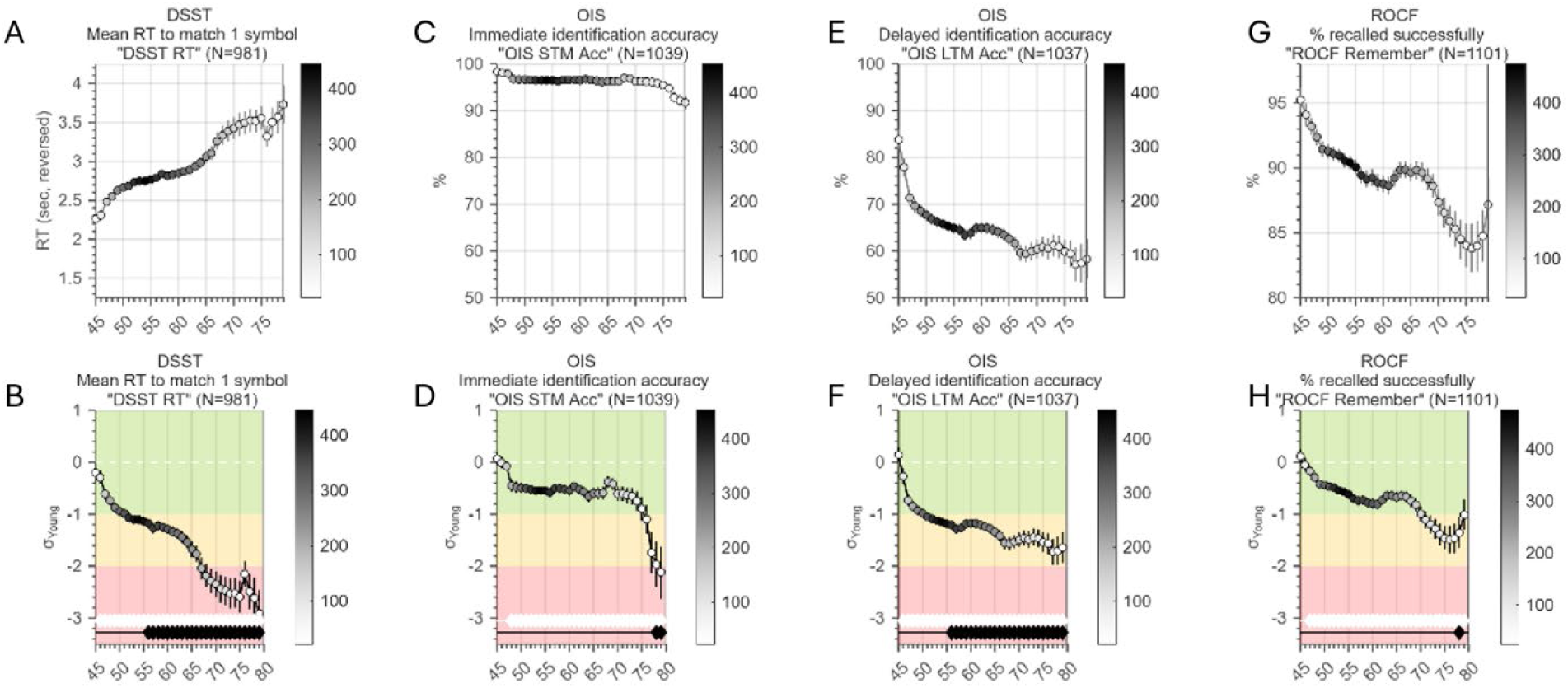
Age trajectories of OCTAL metrics in a healthy population (Study 2, N = 1,109). The top panel displays raw scores for the DSST (A), OIS Short-Term (C) and Long-Term (E) recall, and ROCF memory (G) across ages 45–80 years. Curves are smoothed with a Gaussian kernel (bandwidth = 2.5 years). Marker colour encodes the number of observations contributing to each point, as shown by the colour bar on the right. The bottom panel presents the same metrics normalised to the young-adult reference group (18–39 years). The scale demarcates the normative range (±1 SD, green zone), moderate deviation (1–2 SD, orange zone), and marked impairment (≥2 SD, red zone). Reaction-time variables—such as DSST mean RT in panel A—are sign-reversed so that increasingly negative z-scores indicate greater executive decline. White diamonds along the horizontal white guideline mark age groups that differ significantly from the young-adult mean, whereas black diamonds atop black stems indicate significant deviation from –1 SD (i.e., the boundary between the green and orange zones). See **Supplementary Figure 5** for more metrics.

For memory, immediate identification accuracy (“which object was in this photo?” in the Object-in-Scene task) remained near ceiling into late adulthood (**Figure 2C**) and stayed within the normative band until the late seventies (**Figure 2D**). When memory for the same objects was tested after a brief delay, however, accuracy declined sharply from the mid-fifties onwards (**Figure 2E**). Performance on the Oxford Memory Test (OMT, **Supplementary Figure 5I-J**) and delayed recall of the Rey-Osterrieth Complex Figure (**Figure 2G-H**) also remained within the normative range until the late seventies.

To compare rates of change across domains, we fitted straight lines to the Gaussian-smoothed, age-weighted means of each metric and ranked the resulting slopes (**Figure 3**). The steepest decline was observed for DSST reaction time (–0.073 z-units year⁻¹, R² = 0.95), closely matched by the ROCF copy score (–0.070). Slopes for ROCF immediate recall (–0.035) and OIS delayed identification (–0.031) were roughly half as steep. Full slope estimates and R² values are provided in **Supplementary Table 5** and the individual fit for each metric can be found in **Supplementary Figure 6**.

**Figure 3:**
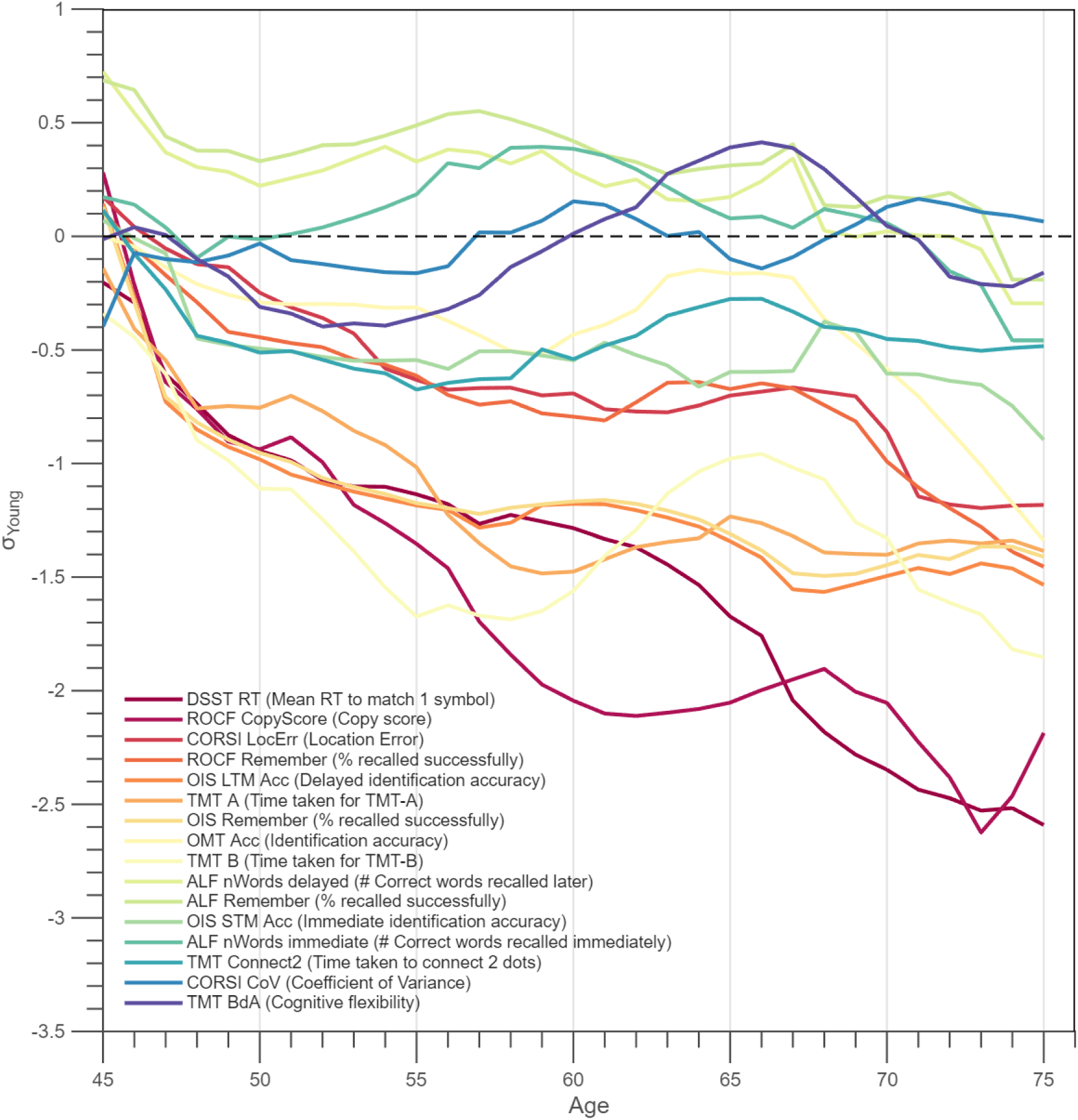
Year-by-year age-related cognitive decline across OCTAL metrics. Lines correspond to the curves from the bottom panel of Figure 2, here combined into a single plot. Metrics are colour-coded and ordered by the magnitude of their negative age slope: warmer (redder) lines denote steeper decline from 45 to 75 years (larger annual reduction in z-score), whereas cooler (bluer) lines denote shallower decline. The y-axis shows performance in standard deviations relative to the mean of the young-adult reference group from Study 1. See Supplementary Table 5 for the slope values and R square of the linear fit (see Methods for details).

Verbal memory showed minimal age effects. In the 12-word list task (abbreviated ALF), immediate recall declined by only –0.009 z-units year⁻¹ and delayed recall by –0.020 z-units year⁻¹. Each list contained one word from twelve semantic categories, pseudo-randomised across 100 sets; participants studied a set to at least 50 % criterion (i.e., they had to remember at least 6 words), then attempted free recall after a 20-minute interval, with only exact spellings scored as correct. Adults aged 45 + actually recalled marginally more words than younger participants (∼9 items on average), and despite smaller samples in the late sixties and seventies, most scores remained within the normative range and did not differ significantly from the lower threshold (**Figure 3**).

### Interrelationships between cognitive domains

A key finding from this large healthy cohort (N=1,109) was the surprisingly modest strength of the correlations between most cognitive metrics, with the majority of Spearman’s ρ values falling below 0.4 (**Figure 4**, or see the full correlation matrix with unadjusted p values in **Supplementary Figure 7**). This suggests that the OCTAL battery measures a set of relatively distinct cognitive processes, rather than a single latent ‘g’ factor. To understand the structure of these relationships at different levels of granularity, we employed two complementary analytical approaches.

**Figure 4.**
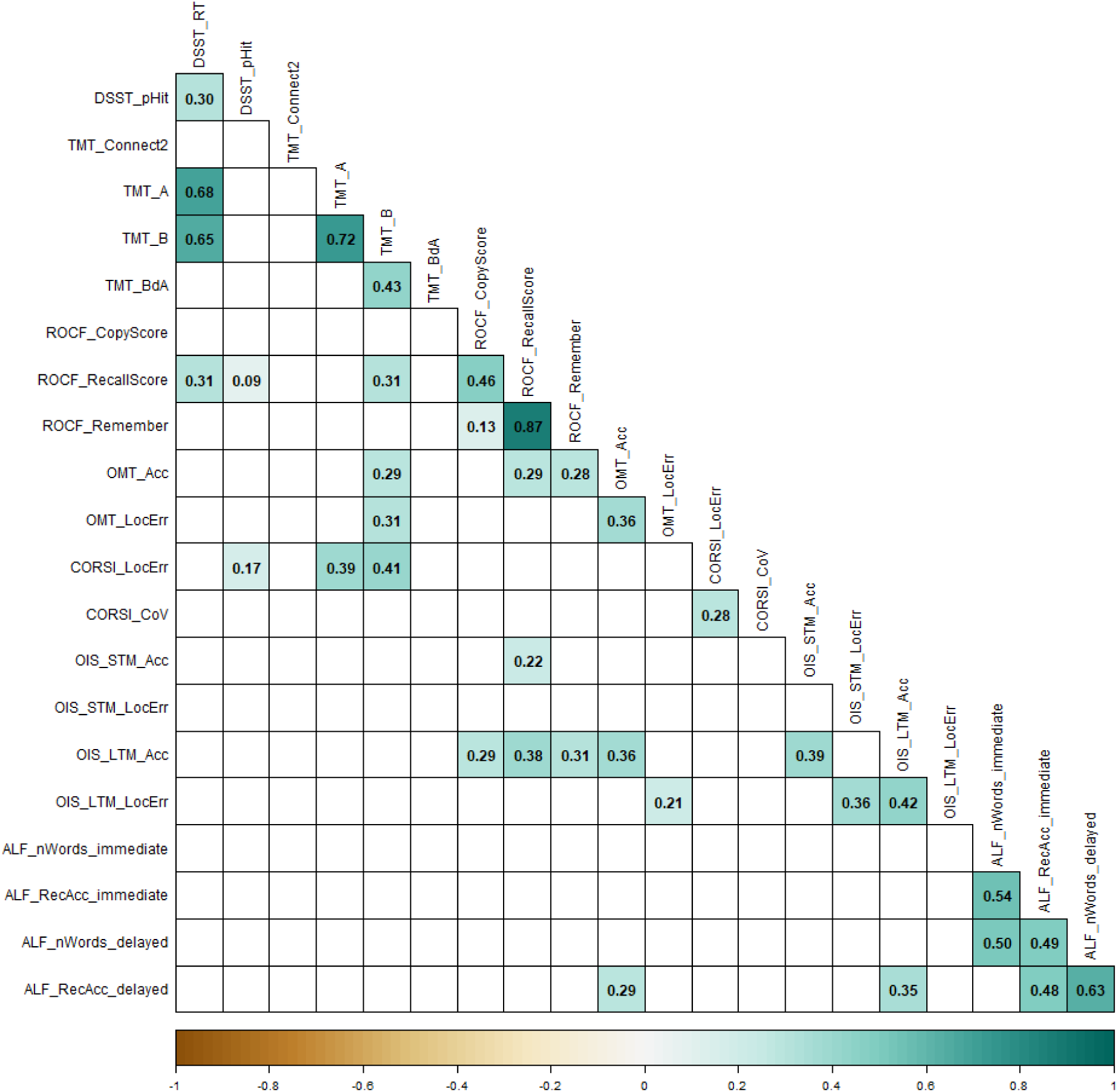
Metric-by-metric correlations among OCTAL indices in healthy adults (Study 2). The matrix displays Spearman coefficients (ρ) for all pairwise comparisons; only correlations that remain significant after Bonferroni correction (p < 0.05) are shown. All metrics are directionally aligned so that lower scores indicate greater cognitive impairment. Square colour, mapped to the accompanying colour bar, encodes the magnitude and sign of ρ, and each cell is labelled with its exact value. Full correlation coefficients and unadjusted p values are reported in **Supplementary Figure 6**.

First, a high-level exploratory factor analysis (EFA) was used to identify the main organising principles of the cognitive data. This analysis revealed a clear and interpretable two-factor structure (using varimax rotation) corresponding to the classical domains of **(1) Memory** and **(2) Executive Function** (Supplementary Figure 4). The analysis confirmed the suitability of the data for factorisation (KMO = 0.68; Bartlett’s Test: χ² = 539.14, p < 0.001) and the sufficiency of a two-factor solution (χ² = 235.54, df = 134, p < 0.001). To allow for inter-factor correlations, we repeated the analysis using promax rotation, which yielded a similar 2-factor structure. Memory and executive function were weakly negatively correlated (factor correlation = −0.2), possibly reflecting individual differences in cognitive strategies that prioritize either accuracy or speed, reinforcing their distinctness.

Second, to investigate the finer-grained relationships within and between these broad domains, we constructed a weighted, undirected network from 25 key metrics and applied a single-pass Louvain-style modularity maximisation algorithm (**Figure 5**, see Methods for details). This method does not require strong correlations but identifies communities of nodes that are more densely interconnected with each other than with the rest of the network. The analysis partitioned the network into six distinct modules. These modules appear to reflect both task-specific effects and broader cognitive functions:

1. **Executive function**: This module’s central node was TMT-A, and it included other members such as DSST_RT, DSST Accuracy, and TMT-Connect2Dots.
2. **Completion time**: Centred on the ROCF copy duration, this two-node module also contained the ROCF recall duration, likely reflecting task strategy and motor speed.
3. **CORSI module**: This module was highly task-specific, consisting only of the CORSI localisation error (the central node) and its coefficient of variation.
4. **Short-term memory**: This module contained all metrics from the OMT, with OMT_Guessing as the central node.
5. **Visuospatial precision**: With OIS long-term memory localisation error as its central node, this module primarily grouped metrics of spatial accuracy, including the ROCF Copy Score and OIS short-term memory localisation error. Interestingly, it also contained the TMT B/A ratio, a measure of cognitive flexibility, suggesting a link between executive control and the performance of precise visuospatial tasks.
6. **Episodic memory**: This was a large, cross-modal memory module with delayed verbal free recall (ALF_nWords_delayed) as its central node. It connected the other verbal memory (ALF) metrics with visuospatial memory retention (ROCF_Remember) and both short- and long-term object identification accuracy (OIS_STM_Acc and OIS_LTM_Acc).

**Figure 5:**
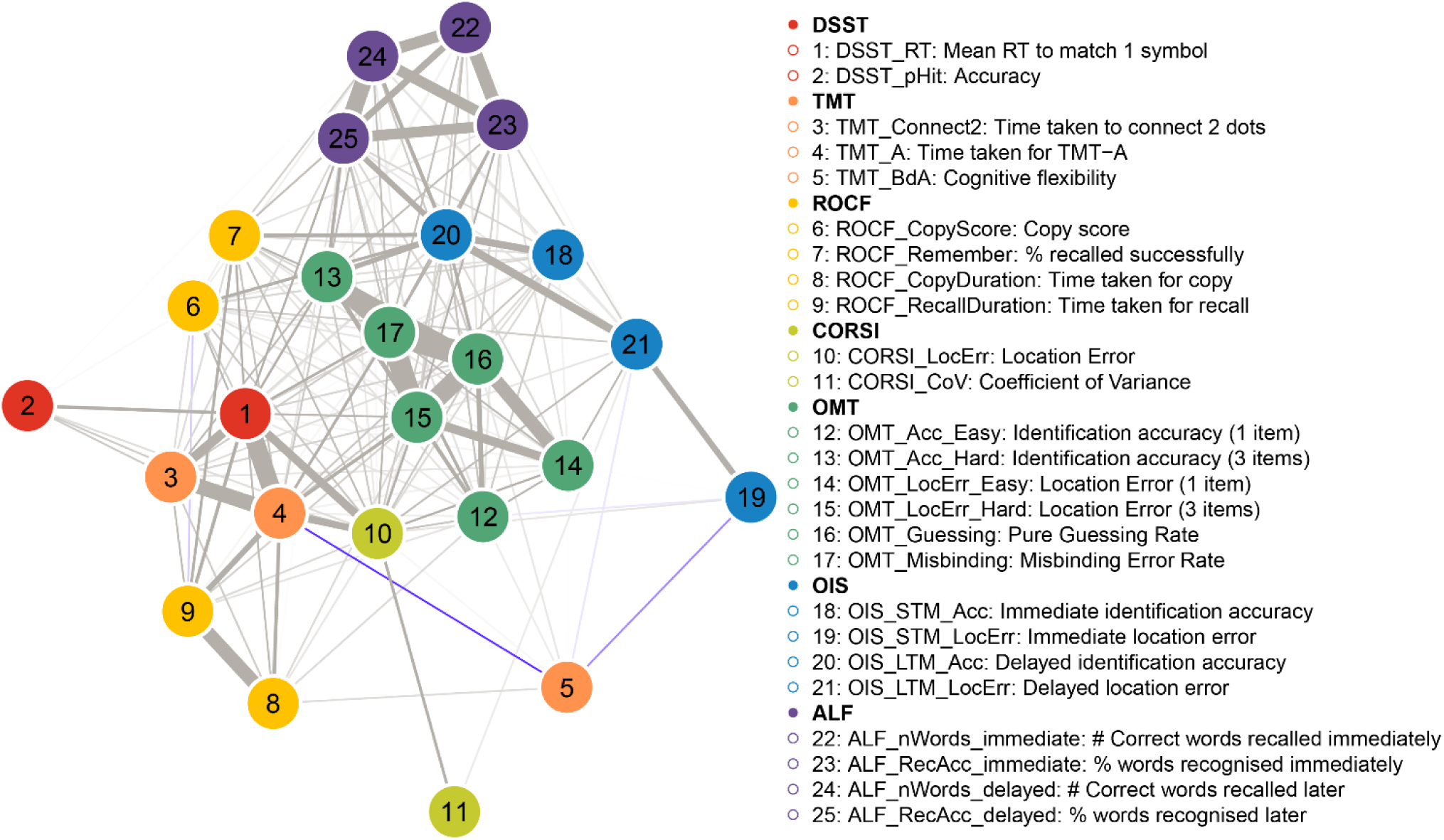
Network plot of relationships among OCTAL key metrics in the healthy population (Study 2). Each node represents one metric and is colour-coded by task (legend ordered by task). Edge thickness and inter-node distance scale with the absolute Spearman correlation coefficient (ρ): shorter, thicker edges denote stronger associations. Edges are displayed only when |ρ| ≥ 0.10. All metrics are directionally aligned so that lower scores signify greater cognitive impairment. Grey edges indicate positive correlations, where better performance on metric A accompanies better performance on metric B. Unsurprisingly, most relationships are positive. Only two edges are negative (highlighted in violet): between metric 5 (TMT cognitive flexibility, defined as TMT-B ⁄ TMT-A) and metric 4 (TMT-A completion time), and between metric 5 and metric 19 (OIS short term location error). Full correlation coefficients and unadjusted p values are reported in **Supplementary Figure 6**. The correlation matrix with multiple comparison correction applied is in Figure 5.

To confirm the stability and robustness of the detected community structure, we ran the Louvain algorithm for 10,000 iterations. The 6-module solution demonstrated remarkable robustness, being obtained in 83.99% of these independent runs. This high consistency, despite the algorithm’s internal randomisation (due to arbitrary node ordering), indicates that the 6-module partition is a highly stable and recurring local maximum in the modularity landscape of our cognitive network. The modularity Q for this dominant solution was consistently low (Q=0.13±0.00), suggesting that while clear communities exist, the inter-module connections are not vastly sparser than expected by chance for this specific network, or that the algorithm tends towards a coarse-grained partitioning given the correlation strengths.

Taken together, these analyses resolve the apparent paradox of finding structured modules despite weak inter-correlations. The EFA reveals two broad, classical cognitive domains, while the network analysis demonstrates that these domains are not monolithic. Instead, they are composed of multiple, weakly correlated sub-components. This finding is a key strength of the OCTAL battery. It indicates that the individual tasks provide high-resolution, non-redundant information, enabling fine-grained cognitive phenotyping.

## Study 3: Does remote testing performance align with in-person standard cognitive assessment?

194 individuals —spanning 97 cognitively healthy controls, 54 with early Alzheimer’s disease dementia, 11 with mild cognitive impairment and 32 with subjective cognitive decline—completed OCTAL remotely and the Addenbrooke Cognitive Examination-III (abbreviated as ACE-III below) during an in-person visit, with test order pseudo-balanced across the cohort. Spearman rank correlations revealed a strong association between OCTAL metrics and the ACE-III total score (**Table 3**). The metrics from the wordlist recall task (coded as ALF) showed the highest concordance (ρ = 0.61–0.74). After Bonferroni correction, three variables did not reach significance (*α* = 0.05): Digit-Symbol hit rate (ρ = 0.15), CORSI Coefficient of variance in localisation accuracy (CORSI_CoV, ρ = 0.06) and Object-in-Scene short-term localisation accuracy (OIS_STM_LocErr, ρ = 0.13).

**Table 3:** Correlations linking ACE-III and OCTAL metrics in Study 3 (N = 194). All OCTAL variables were normalised against age-matched reference data; ACE-III scores are presented as raw scores, consistent with standard practice. Spearman correlation matrix relating the ACE-III total score and the subscores to OCTAL metrics. All p values have been adjusted for multiple comparison.

| OCTAL Metrics | ACE-III Total |  | ACE-III Attention |  | ACE-III Memory |  | ACE-III Visuospatial |  | ACE-III Fluency |  | ACE-III Language |  |
| --- | --- | --- | --- | --- | --- | --- | --- | --- | --- | --- | --- | --- |
| | $\rho$ | adjusted $p$ | $\rho$ | adjusted $p$ | $\rho$ | adjusted $p$ | $\rho$ | adjusted $p$ | $\rho$ | adjusted $p$ | $\rho$ | adjusted $p$ |
| DSST_RT | 0.55 | <0.001 | 0.44 | <0.001 | 0.50 | <0.001 | 0.48 | <0.001 | 0.44 | <0.001 | 0.33 | 0.002 |
| DSST_pHit | 0.15 | 1.000 | 0.22 | 0.236 | 0.16 | 1.000 | 0.20 | 0.433 | 0.06 | 1.000 | 0.04 | 1.000 |
| TMT_Connect 2 | 0.49 | <0.001 | 0.38 | <0.001 | 0.42 | <0.001 | 0.33 | <0.001 | 0.41 | <0.001 | 0.33 | <0.001 |
| TMT_A | 0.59 | <0.001 | 0.51 | <0.001 | 0.59 | <0.001 | 0.38 | <0.001 | 0.50 | <0.001 | 0.34 | <0.001 |
| TMT_BdA | 0.50 | <0.001 | 0.47 | <0.001 | 0.47 | <0.001 | 0.41 | <0.001 | 0.41 | <0.001 | 0.39 | <0.001 |
| ROCF_CopyScore | 0.25 | 0.015 | 0.28 | 0.003 | 0.24 | 0.023 | 0.27 | 0.005 | 0.20 | 0.139 | 0.09 | 1.000 |
| ROCF_Remember | 0.41 | <0.001 | 0.31 | 0.001 | 0.38 | <0.001 | 0.26 | 0.012 | 0.36 | <0.001 | 0.28 | 0.005 |
| CORSI_LocErr | 0.46 | <0.001 | 0.38 | <0.001 | 0.42 | <0.001 | 0.40 | <0.001 | 0.35 | <0.001 | 0.31 | 0.001 |
| CORSI_CoV | 0.06 | 1.000 | -0.02 | 1.000 | 0.06 | 1.000 | -0.02 | 1.000 | 0.00 | 1.000 | 0.02 | 1.000 |
| OIS_STM_Acc | 0.48 | <0.001 | 0.43 | <0.001 | 0.40 | <0.001 | 0.36 | <0.001 | 0.46 | <0.001 | 0.33 | <0.001 |
| OIS_STM_LocErr | 0.13 | 1.000 | 0.05 | 1.000 | 0.14 | 1.000 | 0.00 | 1.000 | 0.09 | 1.000 | 0.10 | 1.000 |
| OIS_LTM_Acc | 0.58 | <0.001 | 0.41 | <0.001 | 0.55 | <0.001 | 0.39 | <0.001 | 0.50 | <0.001 | 0.42 | <0.001 |
| OIS_LTM_LocErr | 0.28 | 0.005 | 0.14 | 1.000 | 0.25 | 0.022 | 0.23 | 0.067 | 0.19 | 0.293 | 0.17 | 0.495 |
| OMT_Acc | 0.47 | <0.001 | 0.42 | <0.001 | 0.41 | <0.001 | 0.37 | <0.001 | 0.45 | <0.001 | 0.35 | 0.002 |
| OMT_LocErr | 0.46 | <0.001 | 0.31 | 0.011 | 0.46 | <0.001 | 0.32 | 0.009 | 0.41 | <0.001 | 0.33 | 0.007 |
| ALF_nWords_immediate | 0.74 | <0.001 | 0.41 | 0.029 | 0.76 | <0.001 | 0.53 | <0.001 | 0.73 | <0.001 | 0.50 | 0.002 |
| ALF_RecAcc_immediate | 0.61 | <0.001 | 0.27 | 0.829 | 0.75 | <0.001 | 0.44 | 0.011 | 0.59 | <0.001 | 0.50 | 0.002 |
| ALF_nWords_delayed | 0.67 | <0.001 | 0.35 | 0.147 | 0.69 | <0.001 | 0.55 | <0.001 | 0.60 | <0.001 | 0.46 | 0.007 |
| ALF_RecAcc_delayed | 0.66 | <0.001 | 0.41 | 0.033 | 0.71 | <0.001 | 0.50 | 0.001 | 0.61 | <0.001 | 0.46 | 0.006 |

Because ACE-III subscores are highly inter-correlated (mean inter-correlation ρ = 0.74; **Supplementary Table 6**), partial correlations were computed while controlling for the remaining four domains (**Figure 6**). Trail-Making Test part A, Trail-Making cognitive flexibility (ratio between time taken to complete trails B and A) and object identification accuracy in immediate recall of Object-in-Scene memory task (short-term memory) were uniquely associated with the ACE-III Attention subscore, whereas the immediate and delayed wordlist recall mapped specifically to ACE-III Memory. Identification accuracy in OMT task and both the short- and long-term phases of OIS showed moderate, selective correlations with ACE-III Fluency. No OCTAL metric exhibited a domain-specific relation with ACE-III Visuospatial or Language.

**Figure 6:**
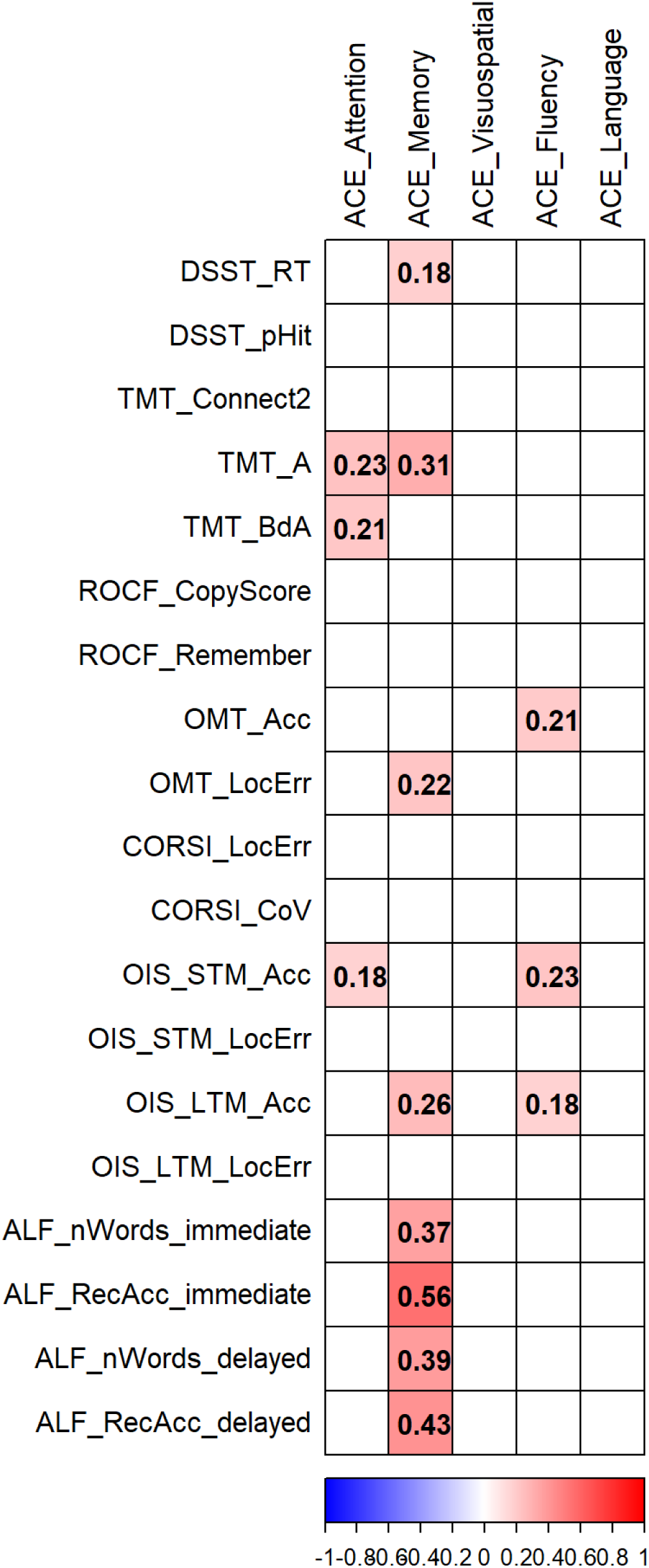
Partial correlation matrix linking ACE-III subscores and OCTAL metrics in Study 3 (N=194). All OCTAL variables were normalised against age-matched reference data; ACE-III scores are presented as raw scores, consistent with standard practice. Partial correlations between each ACE-III subscore and the OCTAL metrics, calculated while regressing out the remaining four subscores (e.g. for ACE-Attention, memory, visuospatial, fluency, and language were removed as covariates).

A machine learning–based feature-ranking algorithm identified ROCF immediate recall score, Object-in-Scene object identification accuracy in delayed recall (long term memory) and time taken to complete TMT-A as the three most informative predictors of ACE-III-defined cognitive impairment (cutoff = 88, **Figure 7A**). Each of these top metrics achieved excellent discrimination in isolation, with area-under-the-curve (AUC) values ≥ 0.88 (**Figure 7B**) and optimal raw-score thresholds of: ROCF immediate recall score < 68.9 % (AUC = 0.91, sensitivity = 79.1 %, specificity = 89.9 %), Object-in-Scene object identification accuracy < 45 % (AUC = 0.90, sensitivity = 90.0 %, specificity = 83.3 %) and Trail-Making Test part A > 32.8 s (AUC = 0.88, sensitivity = 85.0 %, specificity = 84.9 %). Performance was equivalent when z-scores were substituted for raw scores (ROCF recall score: < −1.0 SD, AUC = 0.89, Sensitivity = 81.4%, specificity = 86.0%; Object-in-Scene identification accuracy: <-0.8 SD (AUC=0.89, Sensitivity = 87.5% and specificity = 83.3%; TMT A <-0.9 SD (AUC=0.88, Sensitivity =78.9% and specificity = 84.7%). Although not ranked as top, the 2-minute executive function task —the Digit-Symbol Substitution Test— alone also produced strong classification (AUC = 0.84; **Figure 7B**).

**Figure 7:**
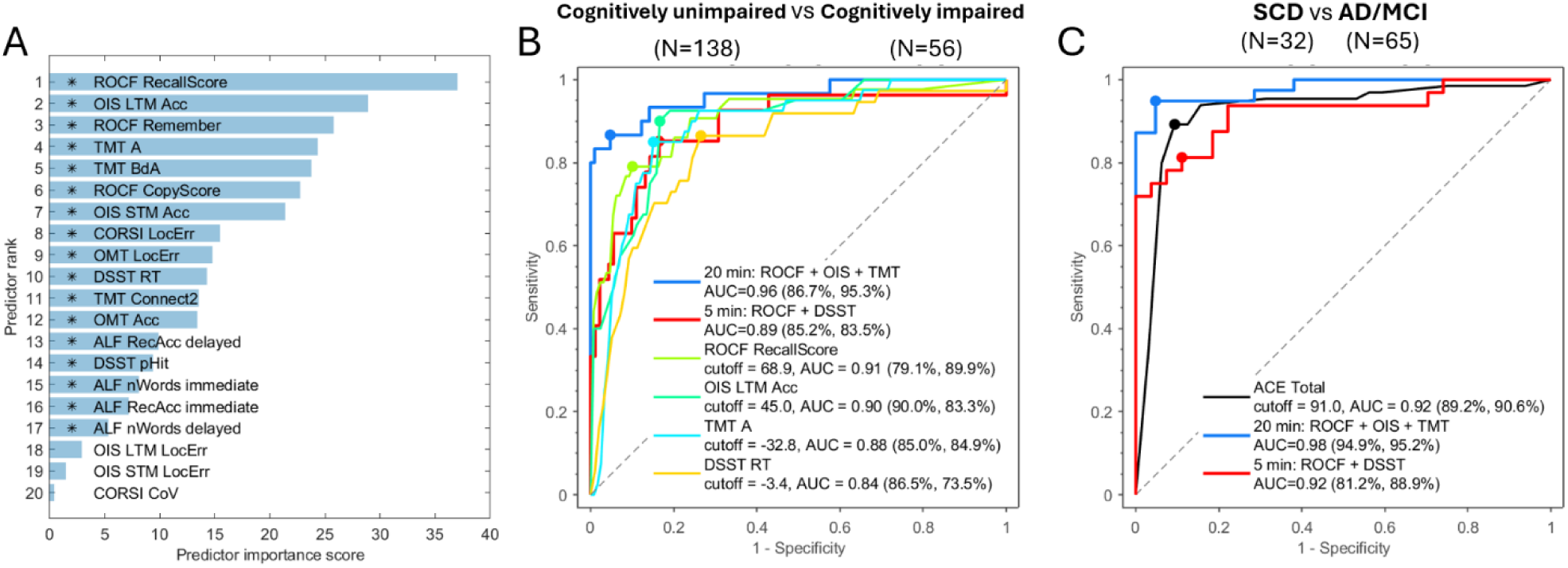
OCTAL performance in detecting ACE-III–defined cognitive impairment and differentiating clinical cohorts. (A) Univariate χ² feature ranking of OCTAL metrics for ACE-III defined cognitive impairment (i.e., ACE-III-Total score < 88) in study 3. Of 194 participants, 138 scored ≥ 88 (normal cognition) and 56 scored < 88 (cognitive impairment), irrespective of diagnosis or subjective symptoms. Higher importance scores denote greater predictive value; metrics are ordered accordingly, and * marks statistical significance. Scores were converted to P-values using 𝑃𝑃 = 𝑒𝑒^−score^. (B) Receiver operating characteristic (ROC) curves illustrate the classification accuracy for cognitive impairment. The area under the curve (AUC) values are shown for each model. For single metric models, the estimated optimal cutoff is shown. The percentage values in brackets indicate sensitivity and specificity, respectively. (C) ROC analysis comparing OCTAL subsets with ACE-III Total for distinguishing participants with neurodegeneration (Alzheimer’s disease, n = 54; mild cognitive impairment, n = 11; total = 65) from those with subjective cognitive decline but no objective impairment (n = 32). DeLong’s test shows the 20-min OCTAL subset marginally outperforms ACE-III Total (p = 0.04), whereas the 5-min subset (red line) yields a comparable AUC.

Combining these top three tasks (ROCF, OIS & TMT), which in total take about 20 minutes of testing, yielded an AUC of 0.96, with sensitivity = 86.7 % and specificity = 95.3 % (see **Figure 7B** and **Supplementary Table 7** for the multiple linear regression model regression model’s weights). When combining ROCF with DSST, which in total should take no more than 5 minutes, this rapid five-minute screening set achieved an AUC of 0.89 with balanced good sensitivity (85.2 %) and specificity (83.5 %).

Can these combinations of OCTAL tasks reliably distinguish people with clinical diagnoses of a neurodegenerative disease from subjective cognitive decline? In a subset of 97 participants (65 with Alzheimer’s disease dementia or mild cognitive impairment; 32 with subjective cognitive decline), the 20-minute OCTAL composite yielded an AUC = 0.98, significantly outperforming the ACE-III total score (AUC = 0.92; DeLong p = 0.042; see **Figure 7C**). A compressed 5-min OCTAL composite matched the accuracy of ACE-III (AUC = 0.92 for both; p = 0.40).

To further probe its clinical utility, we used multiple linear regression to determine if performance on the 20-minute OCTAL battery (comprising the ROCF, OIS, and TMT) could predict scores on the in-person ACE-III. The model proved to be a strong predictor of the ACE-III total score (r = 0.83, p < .001). High predictive accuracy was also achieved for the Attention, Memory, Visuospatial, and Fluency subscores, with correlations ranging from r = 0.75 to 0.80 (all p < .001). A significant, though more moderate, correlation was found for the Language subscore (r = 0.57, p < .001); this may be partly explained by the high degree of inter-correlation between the ACE-III subscores themselves, where performance on the language domain is also associated with other cognitive functions. The close correspondence between OCTAL-predicted and actual clinician-administered scores for all domains is illustrated in **Figure 8**.

**Figure 8:**
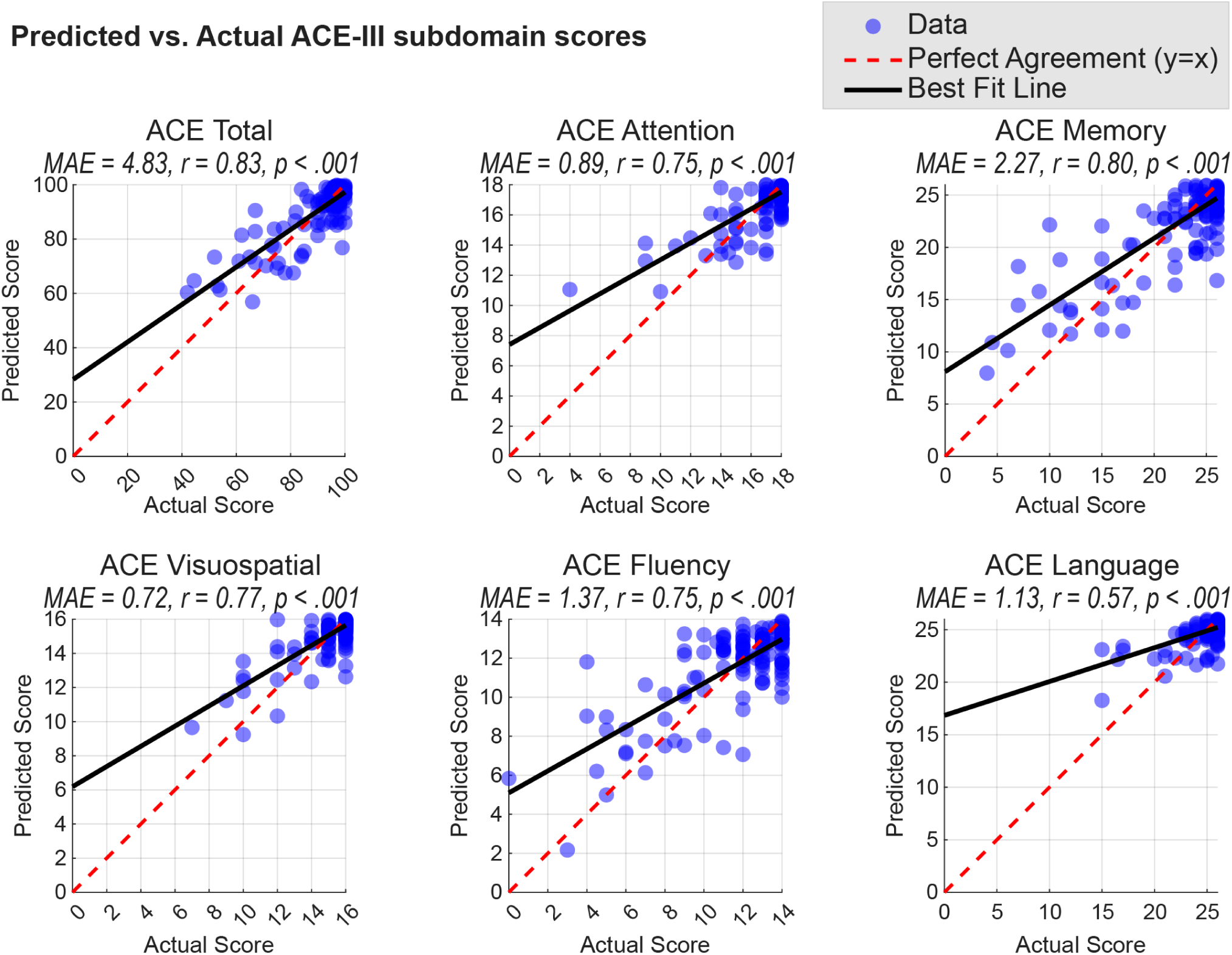
Predicting ACE-III subscores from remote OCTAL performance in Study 3. Each scatter plot compares scores on the ACE-III (total and subdomains) predicted by a multiple linear regression model against the actual, clinician-administered scores. The models used performance on the ROCF, OIS, and TMT tasks as predictors. Blue circles represent individual participants. The solid black line shows the linear best fit, while the dashed red line indicates perfect agreement (y=x). Each panel is annotated with the Mean Absolute Error (MAE) and the Pearson correlation coefficient (r) for that model.

Collectively, these results demonstrate that remote OCTAL assessment aligns closely with in-person ACE-III performance, provides domain-specific cognitive insights and accurately discriminates pathological from non-pathological ageing within brief testing times.

## Study 4: Six-month test–retest reliability across three visits

A total of 118 participants—59 cognitively healthy controls, 23 individuals with subjective cognitive decline, five with mild cognitive impairment and 31 with Alzheimer’s disease dementia—completed three OCTAL assessments at baseline, three months and six months. These participants additionally undertook the paper-based ACE-III using the alternate A, B and C forms. Test–retest reliability was quantified with the intraclass correlation coefficient (ICC) defined by McGraw and Wong,^19^ which was calculated on the continuous performance scores across the three visits. Following the interpretative framework of Koo and Li^20^, values below 0.50 were deemed poor, 0.50–0.74 as moderate, 0.75–0.90 as good and those exceeding 0.90 as excellent. All coefficients, ranked from lowest to highest, are displayed in **Figure 9**, with their 95 % confidence intervals and F-statistics reported in **Supplementary Table 7**.

**Figure 9:**
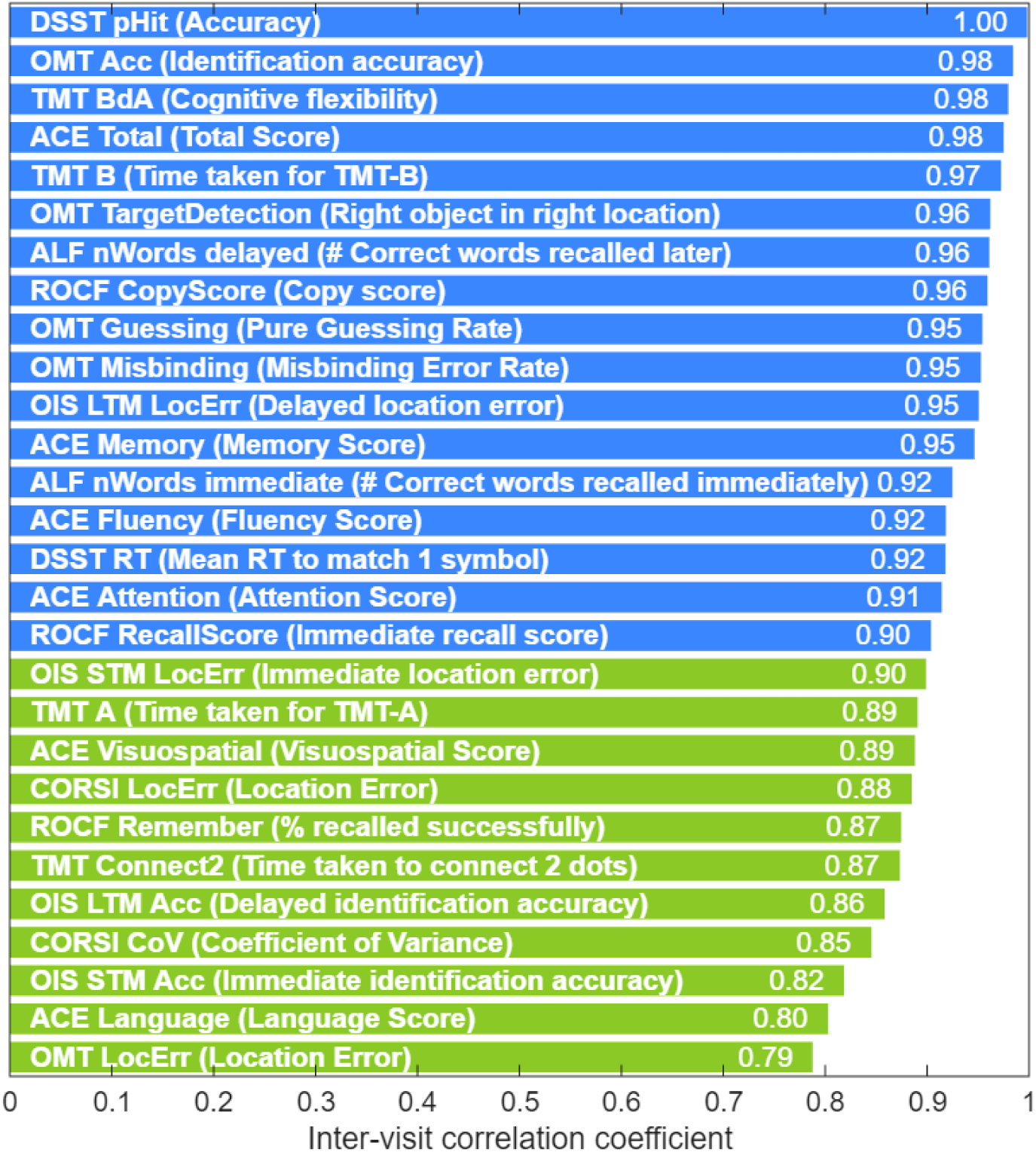
Test–retest reliability ranking. Intraclass correlation coefficients (ICCs) are presented for all OCTAL metrics across three visits, alongside the ACE-III total and subdomain scores. ICCs were classified as poor (< 0.50), moderate (0.50–0.74), good (0.75–0.90), or excellent (> 0.90). Metrics with good reliability are coloured green; those with excellent reliability are coloured blue.

The ACE-III total score demonstrated excellent stability over three tests (ICC = 0.98, 95 % CI 0.97–0.98). Domain scores were likewise robust, with Memory 0.95, Fluency 0.92, Attention 0.91, Visuospatial 0.89 and Language 0.80; each coefficient significantly exceeded the 0.75 criterion for good reliability (p < 0.001). Many OCTAL-derived metrics surpassed these benchmarks and, in several instances, outperformed the ACE-III total score itself. The most stable indices were the Digit Symbol Substitution Test hit rate, Object-Memory Test identification accuracy and the cognitive-flexibility ratio from the Trail-Making Test (TMT-B time divided by TMT-A time), all with ICCs ≥ 0.98. The least stable measure, the Object-Memory Test location error, still achieved an ICC above 0.75, denoting good reliability. these findings confirm that every principal OCTAL metric possesses good to excellent six-month test–retest reliability, comparable with—and in several instances superior to—the ACE-III and its subscores.

## Discussion

Here, we introduced the Oxford Cognitive Testing Portal (OCTAL), a remote, browser-based battery that measures memory, attention, visuospatial and executive functions (**Figure 1**). Across four studies involving 2,595 adults—healthy controls and individuals with Alzheimer’s disease dementia, mild cognitive impairment or subjective cognitive decline—OCTAL proved accessible across the adult lifespan, reliable without clinician supervision, and fully functional once translated on standard hardware. Crucially, patients with AD dementia and MCI completed OCTAL remotely without any clinician present, demonstrating its feasibility in cognitively impaired populations.

OCTAL builds upon a growing ecosystem of digital cognitive assessment tools, as reviewed recently in ^13^, including established platforms like CANTAB,^21^ the Cogstate Brief Battery,^22^ and Cognitron^18^. A critical challenge for remote digital assessments, highlighted in large-scale studies with over 100,000 participants, is the difficulty in maintaining participant engagement.^23^ With this challenge in mind, OCTAL was uniquely co-designed from the ground up by clinicians, cognitive neuroscientists, and patients themselves. This collaborative approach ensures that the platform not only meets clinical needs and scientific standards, but is also engaging for the patients with cognitive impairment. A key advantage that distinguishes OCTAL is its open, modular architecture hosted on Pavlovia, a platform widely used by cognitive and psychological researchers.^24–26^ This architecture facilitates straightforward integration with other web-based tools, such as Cognitron, and allows for the seamless incorporation of novel cognitive tests as they are developed. This inherent flexibility makes OCTAL a uniquely sustainable and evolvable tool for the research community.

In the present study, the visuoconstructive recall (ROCF), delayed object–scene recognition (OIS) and visual processing speed (TMT-A) emerged as the strongest predictors of ACE-III-defined cognitive impairment (a standard face-to-face cognitive screening test) in the machine-learning analyses of 194 individuals (**Figure 7A**). These top three tasks, taking 20 minutes in total, showed excellent performance for detecting cognitive impairment defined by an ACE-III cutoff of < 88 (AUC = 0.96). The combination of ROCF and DSST also performed well (AUC = 0.89) (**Figure 7B**). Furthermore, the 20-minute OCTAL subset outperformed the in-person ACE-III in distinguishing AD or MCI from subjective complaints (SCD), while an ultra-brief five-minute version (ROCF + DSST) matched ACE-III accuracy with markedly lower burden (see **Figure 7C**). Furthermore, the modelling demonstrated that remote OCTAL performance could predict the specific scores for each ACE-III cognitive domain, showing a strong correlation with the actual clinician-administered scores for memory, attention, fluency, and visuospatial functions (**Figure 8**). This highlights that OCTAL not only provides a valid global measure of cognition but also offers a granular, domain-specific profile that aligns closely with traditional, in-person assessments.

OCTAL provides very sensitive domain-specific measures of executive functions and memory. This is demonstrated in Study 2’s investigation of healthy cognitive ageing, where OCTAL quantifies domain-specific change across five decades from mid to late life. Distinct cognitive ageing trajectories were quantified, with OCTAL revealing that executive functions decline from the mid-forties while memory remains within young-adult normative ranges until the mid-seventies. Long-term object–scene memory showed intermediate decline at ≈ 55 years (**Figure 2F**), emphasising the value of granular digital metrics over composite scores.

Performance remained stable over six months (Study 4, intraclass correlation coefficients ≥ 0.79), confirming robust repeatability. One possible explanation for self-administered OCTAL’s advantage over clinician-administered ACE-III is its avoidance of ceiling effects, which provides greater granularity in high-functioning individuals. Other factors may also contribute. For instance, the digital, automated nature of OCTAL eliminates variability in test administration and scoring that can occur with a pen-and-paper test administered by different clinicians. Furthermore, the millisecond-level reaction time data captured by OCTAL offers a level of precision in measuring processing speed and executive function that is not possible with manually timed, observation-based assessments like the ACE-III.

A further significant advantage of this digital, automated nature is the ability to embed objective performance validity checks. While in the present study we used these checks primarily to flag low-engagement data for exclusion, they offer a much richer opportunity. Performance on these embedded attention probes could be synthesised into a quantitative ‘engagement index’. For a clinician, such an index would provide a formal measure of test validity, replacing the subjective ‘soft features’—such as observing a patient’s effort or hesitation—relied upon during in-person assessments to gauge whether a cognitive score is a true reflection of the individual’s ability. This data-driven approach to measuring engagement is a key step towards more reliable and nuanced remote cognitive assessment.

Importantly, device heterogeneity did not diminish performance, as OCTAL ran reliably on any standard hardware without requiring a uniform screen size or input method. Most OCTAL metrics—such as reaction time and spatial-error indices—were unaffected by screen size or aspect ratio (**Supplementary Figures 2–3**), confirming its suitability for deployment on heterogeneous devices without formal standardisation. Where device effects appear (e.g., fine-motor precision in the ROCF copy task), OCTAL mitigates them by expressing memory performance as the recall-to-copy ratio, effectively regressing out motor variance. Expanding normative datasets across device types may further refine this adjustment.

Furthermore, cross-linguistic validation demonstrated equivalent performance between native English and native Chinese speakers once translations were implemented. The core metrics of the OCTAL were practically indistinguishable between the groups, falling within one standard deviation of the normative range (see **Table 1**). A somewhat surprising finding from our cross-cultural comparison was the consistent, albeit variable, latency in response times in the China-based cohort compared to the UK-based one. This was most pronounced for the initial identification response in the OIS task, but a general trend of slower reaction times was observed across tasks. This finding is important because, to our knowledge, detailed examinations of remote testing latency have been largely confined to North American and European internet environments.^24,26^ The unique and complex nature of internet infrastructure and data handling in other regions, such as mainland China, has not been systematically documented in the context of remote cognitive testing. We do not consider this finding to be a (prohibitive) limitation for the use of time-sensitive measures in diverse online environments. Rather, it highlights the critical need for robust experimental designs that can account for and mitigate these baseline differences. One effective strategy is the use of within-task controls and ratio-based metrics. For example, instead of relying on the absolute completion time of TMT-B, the ratio of TMT-B to TMT-A can provide a purer measure of executive function by factoring out baseline psychomotor speed and general system latency. The observation that the most significant latency difference occurred in a rapid-response component of the OIS task, while being less pronounced in tasks with longer decision times, may suggest a larger initial latency at the beginning of a trial. This initial lag becomes a smaller proportion of the total reaction time in more complex tasks. This underscores the importance of moving beyond simply trusting that device and internet differences are negligible. Future research in remote cognitive assessment must prioritise study designs that are either validated across diverse technological landscapes or include internal controls to ensure that the derived cognitive metrics are robust and truly reflect cognitive processes, not environmental or technical confounders. This is an essential step for achieving genuine international scalability and equity in digital cognitive science.

Another example of cultural influence on cognitive performance is the Trail-Making Test Part B (TMT-B). Even all living in UK (ruling out the potential latency confound due to internet infrastructure), healthy individuals who speak and read English fluently took considerably longer to complete this, while there was no difference in TMT-A. This is likely due to the cognitive load of alphabet switching, a finding consistent with previous reports in Korean speakers using the pen-and-paper version.^27^ This finding underscores the long-standing challenge of ensuring the cross-cultural applicability of cognitive screening tools, a need that persists *even for fluent bilinguals*.^28–30^ Although TMT-B is a highly sensitive and reliable predictor of cognitive impairment—surpassing all ACE-III subscores—its reliance on alphanumeric sequencing presents cultural barriers. Furthermore, its increasing duration with greater deficits poses practical challenges. Consequently, we omitted the TMT-B from the core smartphone version of OCTAL (still collecting data) to maintain both sensitivity and international usability. In its place, we incorporated the Digit Symbol Substitution Test (DSST), which also assesses attention and executive function, has a brief administration time of two minutes, and reliably predicts global cognitive impairment.

Together, these findings establish OCTAL as a scalable, user-friendly and psychometrically robust alternative to traditional in-person cognitive assessment. Its domain-specific sensitivity, minimal participant burden and remote deployment render it ideally suited for large-scale cognitive screening, decentralised clinical trials and personalised longitudinal monitoring in precision neurology.

Moreover, remote cognitive testing has the capacity to fundamentally transform clinical-trial design.^10^ By capturing high-frequency measurements, platforms like OCTAL can increase the statistical power of a study, which can in turn reduce the required sample size and associated costs. This approach also enables decentralised clinical trials, expanding participant access and making research in less common conditions more financially viable. Furthermore, by profiling individual cognitive trajectories rather than relying on group-mean effects, this level of precision supports adaptive trial protocols, accelerates ‘go/no-go’ decisions and facilitates truly personalised therapeutic interventions. Beyond clinical trials, these methods can benefit routine clinical services. The availability of rapid, low-burden assessments marks a significant shift away from infrequent, episodic testing that often requires specialist administration. This increased accessibility can streamline patient triage and help to inform more timely and effective care decisions.^13,14^

Remote testing offers unprecedented opportunities for population health. It allows early detection of prodromal decline at scale, rapid assessments completed in under ten minutes and, critically, unified integration with fluid biomarkers for refined risk stratification. The advent of scalable blood-based biomarkers for Alzheimer’s disease necessitates digital tools that can be deployed remotely and longitudinally alongside them. In fact, in our previous study, we demonstrated strong relationships between OCTAL performance and plasma AD biomarkers (phosphorylated tau 181) in a smaller cohort.^31^ Because no specialised hardware is required, OCTAL is accessible to geographically isolated or mobility-restricted individuals, and it supports long-term monitoring at intervals that would be impractical in a clinic setting. By enabling frequent, detailed sampling of cognitive function, OCTAL promises to accelerate precision medicine, enhance trial efficiency and reduce overall cost in both research and clinical practice.

### Limitations

First, we did not assess the impact of colour-vision deficiencies on task performance. Although we deliberately avoided using colour in most tasks (ROCF, DSST, TMT and ALF) and designed OIS stimuli to differ in shape and orientation rather than hue—and varied fractal forms in OMT—participants with colour blindness may still experience slower visual discrimination. In Studies 3 and 4, colour blindness was one of the exclusion criteria. To address this in the future, we will include a brief screening question on colour vision in future studies. Second, we did not log the extent of caregiver assistance. Caregivers were instructed to clarify task instructions but not to perform the tasks for patients themselves. The strong concordance between OCTAL scores and ACE-III performance suggests that this instruction was generally followed, but formal monitoring would strengthen future protocols. Third, testing took place in uncontrolled home environments, without measures to limit external distractions or to standardise breaks between tasks. Although this reflects real-world conditions, it may introduce additional variance that should be quantified in subsequent work. Fourth, the current version of OCTAL does not include dedicated tests of language function, and none of the existing OCTAL tests correlated with the language subscore of the ACE-III. This might reduce the battery’s sensitivity to isolated language deficits, such as those found in primary progressive aphasia. It should be noted, however, that the predominantly language-independent design is also a key strength, increasing the suitability of the battery for diverse populations where language proficiency could act as a confounding factor. Finally, our data were pooled from multiple projects conducted at different time points, so not every participant completed every task; for example, the ALF task has a substantially smaller sample than the others.

## Methods

### Overview of the OCTAL platform

#### Development and technical specifications

The Oxford Cognitive Testing Portal (OCTAL, https://octalportal.com) is a fully remote, browser-based cognitive assessment platform designed for unsupervised administration on participants’ personal devices, including desktops, laptops, and tablets. The platform’s architecture supports a battery of cognitive tasks targeting diverse cognitive domains.

The majority of OCTAL tasks were developed using PsychoPy Builder (PsychoJS v2022.2.4), an open-source software package widely used for creating behavioural, psychological and cognitive experiments.^24,32^ This choice of development environment promotes reproducibility and facilitates modification by the broader research community. To incorporate specific functionalities not natively available in PsychoJS, a custom-built in-house JavaScript library was developed and integrated. A notable exception to the PsychoJS framework is the Rey–Osterrieth Complex Figure (ROCF) task, which was custom-built from the ground up using HTML5 and JavaScript. A detailed discussion of the rationale and copyright considerations for the use of this stimulus is provided in the ‘Ethical and intellectual property considerations’ section. This approach was adopted to accommodate the complex drag-and-drop interactions required for its digital administration. This combination of standardised and bespoke development tools allowed for both efficient creation of common task elements and the flexibility needed for novel psychometric paradigms. The platform’s open and modular architecture, a key design principle, facilitates straightforward integration with other web-based research tools and supports the future incorporation of new cognitive tests.

#### Hosting, data anonymisation and security

All OCTAL tasks are hosted on Pavlovia.org, operating under the institutional licence of the University of Oxford’s Department of Experimental Psychology. Pavlovia.org is an established platform for hosting online psychological experiments, providing robust infrastructure and data management capabilities. Throughout the data collection period detailed in this manuscript, OCTAL was deployed exclusively via this infrastructure. To support wider academic access and enhance scalability for future research, the platform is currently being transitioned to a dedicated server environment, also powered by Pavlovia (https://pavlovia.octalportal.com/).

Participant data are handled with strict adherence to privacy and security protocols. All collected data are fully anonymised; cognitive performance data are stored in an encrypted database, physically and logically separated from any personal identifying information. The Pavlovia.org platform employs comprehensive security measures, including the use of EU-based servers that comply with stringent data protection regulations, such as the General Data Protection Regulation (GDPR). All data transmission occurs over secure HTTPS connections utilizing advanced encryption standards. Furthermore, access to the platform is blocked for browsers that do not support modern encryption technologies, ensuring a high level of data protection during remote assessment. These measures are critical for maintaining participant confidentiality and data integrity, particularly when collecting sensitive cognitive information from diverse populations, including clinical cohorts, in unsupervised remote settings.

#### Usability and accessibility

OCTAL was designed with a strong emphasis on usability and accessibility to ensure broad applicability. All tasks are delivered via standard web browsers, eliminating the need for participants to download or install any software, and no user log-in is required for participation. This approach minimizes barriers to entry and simplifies the participant experience. The platform is compatible with a wide range of common web browsers, including Chrome, Firefox, Safari, and Edge, and is designed to function across various operating systems and devices, such as desktops, laptops, and tablets.

A core design principle was the optimisation of task presentation and interaction to ensure consistent performance and user experience regardless of variations in screen size or device type. This was partly addressed through specific analyses of screen size effects (detailed in Study 1) and the implementation of procedures like card calibration for tasks sensitive to stimulus dimensions (e.g., Oxford Memory Task). Critically, OCTAL was co-designed by a multidisciplinary team comprising clinicians, cognitive neuroscientists, programmers and patients with cognitive impairment. This collaborative, patient-centred design process was instrumental in creating a platform that not only meets rigorous scientific and clinical standards but is also engaging and intuitive for users, including older adults and individuals with cognitive difficulties. The task flow was deliberately optimised to be straightforward for these populations, addressing known challenges related to participant engagement and adherence in remote digital health studies.

#### Task administration and flow

To ensure a smooth and coherent testing experience, a custom JavaScript-based script manages seamless transitions between individual cognitive tasks within the OCTAL. This script automatically redirects participants from one completed task to the next, programmatically preserving a unique, anonymised participant identifier embedded within the URL. This system minimises the cognitive load on participants, minimise the data loss and reduces the likelihood of attrition between tasks.

The method for generating and managing participant links was adapted to different recruitment strategies. For large-scale online recruitment via platforms such as Prolific, the redirection and participant ID tracking were fully automated using Prolific’s integrated participant ID system. For participants recruited from clinical settings (e.g., memory clinics), personalised study links were generated and sent via email following the informed consent process. These links embedded anonymised participant IDs and specific trial or visit identifiers, ensuring correct data attribution. Participants were generally encouraged to complete the entire battery of tasks in a single session to maintain consistency.

For researchers and clinicians, a secure backend interface, managed via GitLab, provided real-time access to monitor data completion and quality. This facility allowed for proactive identification of any technical issues or incomplete submissions. The system was designed to save incomplete data, with provisions for data deletion should a participant choose to withdraw consent, although no such requests were made during the studies reported.

#### Ethical and intellectual property considerations

The intellectual property associated with the Oxford Cognitive Testing Portal (OCTAL) is formally managed by Oxford University Innovation (OUI), under project number 22768. Applications for a free academic licence can be made at: octalportal.com/apply-academic-license

All visual stimuli and assets used within the OCTAL platform were either created for this project, are available in the public domain, or are used with permission from the copyright holders.

A specific consideration relates to our use of the Rey–Osterrieth Complex Figure (ROCF). The stimulus for this task is the figure originally designed by Rey and later standardised by Osterrieth.^33,34^ Our use of this figure is consistent with the principles of fair dealing for non-commercial academic research. Crucially, our novel digital adaptation employs only the figure as a visual stimulus; it does not use or replicate the copyrighted administration, scoring protocols, or manuals associated with the standardised pen-and-paper test. This digital task employs a drag-and-drop construction method, a procedure that is fundamentally different from the traditional pen-and-paper drawing administration. Despite this procedural difference, we have retained the ‘ROCF’ designation to reflect the task’s primary aim: to assess visuospatial constructional ability, the same core cognitive domain targeted by the original test.

A core feature of this task’s architecture is that it is intentionally stimulus-agnostic. This design allows for the future use of alternative, parallel stimuli, which is critical for mitigating practice effects in longitudinal studies where participants are assessed on multiple occasions. **For the specific purpose of this initial validation study, however, we deliberately employed a single, fixed stimulus.** The ROCF was selected for this benchmark role precisely because its well-characterised complexity and extensive documentation in the neuropsychological literature provide a stable and robust reference point. This approach ensures that our validation focuses on the performance of the digital platform itself, rather than introducing confounding variables from a novel or variable stimulus set.

### Task descriptions

The OCTAL comprises seven distinct tasks (**Figure 1**), many adapted from established neuropsychological tests or behavioural paradigms, and optimised for robust remote administration across heterogeneous devices. These tasks are designed to measure various aspects of human cognition, including different forms of memory, attention, processing speed, and executive functions.

#### Trail Making Test (TMT)

The TMT is a widely used neuropsychological test, adapted for online administration in OCTAL to assess attention, processing speed and executive functions, particularly cognitive flexibility.^35,36^

**Procedure:** Participants are instructed to tap or click on on-screen nodes (circled numbers or letters) as rapidly as possible to connect them in a specified sequence. The task includes three conditions:

1. Control Task (’Connect-2-Dots’): This condition consists of two trials. Two circles, labelled ‘1’ and ‘2’, are presented at opposite corners of the screen. Participants connect ‘1’ to ‘2’. This task primarily gauges basic psychomotor speed and click accuracy.
2. TMT-A (Numeric Trails): This condition comprises three trials. Participants connect 25 circled numbers in ascending numerical order (e.g., 1-2-3-…).
3. TMT-B (Alternating Trails): This condition also comprises three trials. Participants connect 25 circled numbers and letters in an alternating sequence (e.g., 1-A-2-B-3-C-…). For TMT-A and TMT-B, each participant is presented with six different trail maps (three for TMT-A, three for TMT-B), which are randomly selected from a pre-generated pool of 100 unique maps. These maps were created using a “divide-and-combine” algorithm to ensure consistent difficulty and spatial distribution.^37^ The use of a large pool of distinct maps helps to minimize practice effects upon repeated administrations.

A demonstration of the TMT is available at: https://octalportal.com/demo-trail-making/.

**Outcome metrics:** The primary metrics derived from the TMT are mean completion times for each condition: `TMT_Connect2Dots` (control), `TMT_A` (numeric), and `TMT_B` (alternating). Additionally, the ratio of `TMT_B` completion time to `TMT_A` completion time (`TMT_B/A` ratio, also referred to as `TMT_BdA`) is calculated as a specific marker of executive function, reflecting cognitive flexibility or task-switching ability. The digital format allows for millisecond-level precision in timing these responses

#### Digit Symbol Substitution Test (DSST)

The DSST is another commonly used measure of executive functions, processing speed, attention, and associative learning, adapted for digital administration within OCTAL.^38^

**Procedure:** A reference key is continuously displayed at the top of the screen, showing nine unique symbols, each paired with a digit from 1 to 9. Below this key, a row of nine randomised symbols is presented. Participants are required to match each symbol in the bottom row with its corresponding digit from the key by clicking on the correct digit (presented as a choice array). Once all nine symbols in the row are answered, the row is refreshed with a new set of nine randomised symbols. Participants are instructed to complete as many correct matches as possible within a fixed time limit of 2 minutes. A countdown in seconds is shown in the middle of the screen. Before the main task, two matched examples are shown, followed by a practice round with seven examples using a fixed practice key with 9 different symbols which do not appear in the main task. A demonstration of the DSST is available at: https://octalportal.com/demo-dsst/

**Stimuli**: To mitigate practice effects, the main task’s symbol-digit key is randomly generated for each session by selecting nine symbols from a pool of 20 distinct symbols. The rows of symbols for matching are generated dynamically. The symbols were purchased from the *Noun Project* (https://thenounproject.com/).

**Outcome metrics**: Performance is quantified by several metrics:

- `DSST_rt`: The mean inter-response time for correct symbol-digit substitutions, calculated after removing outlier inter-response times (those more than 2 standard deviations from the mean of correct inter-response times). This is the key metric of DSST used in this paper.
- `DSST_pHit`: The proportion of correct matches relative to the total number of responses made. This is the key metric of DSST used in this paper.
- `DSST_nCorrectResponse`: The total number of correct symbol-digit matches completed within the 2-minute duration.
- `DSST_nAllResponse`: The total number of responses made by the participant during the task.
- `DSST_totalIdleTime`: The cumulative duration of excessively long inter-response intervals for correct responses (defined as intervals greater than the median correct inter-response interval plus 3 times its standard deviation), reflecting periods of participant inactivity or disengagement during correct performance. This metric is used to exclude participants, not a cognitive measure.

#### Freestyle Corsi Block Task (CORSI)

This task was inspired by the classic Corsi Block Tapping Task, a standard measure of visuospatial short-term memory span.^39^ In the original version, participants were presented with a set of nine identical wooden blocks positioned on a board. Participants were required to point at the blocks in the order they were presented. On OCTAL, the key modification in our “Freestyle” version is that the spatial locations of the stimuli are not fixed across trials or participants, unlike typical computerised Corsi tasks where blocks appear in predefined positions. In an ‘n-location’ trial, a 1-cm wide red dot appears at a random location on the screen. After 1 second, it disappears. If the sequence length (n) is greater than 1, the dot then reappears at another random location for 1 second. This process repeats ‘n’ times, with sequence lengths tested up to 3 items. After the entire sequence of ‘n’ dots has been presented, there is a 1-second pause. Following this, the participant is instructed to reproduce the sequence by clicking on the screen at the locations where each dot had appeared, in the correct temporal order. The task is divided into three blocks. Each block consists of five trials of a specific sequence length (’n-location sequence’). A demonstration of the CORSI is available at: https://octalportal.com/demo-corsi/

##### Outcome metrics

- `CORSI_LocErr`: The mean localisation error, calculated as the average Euclidean distance between the participant’s clicked response location and the actual target location for each item in the sequence, averaged across all trials and sequence lengths.
- `CORSI_CoV`: The between-trial coefficient of variation of the localisation error, providing a measure of response consistency.

#### Oxford Memory Task (OMT)

The OMT is a visual short-term memory task based on the “What was where?” paradigm, assessing memory for object identity and spatial location. This has been described in detailed in previous publications from our group (Figure 2).^40–42^ This remote online version is shorten based previous data.

**Procedure**: Participants are presented with either one fractal pattern (easy trials) or three fractal patterns (hard trials) simultaneously displayed at various locations on the screen for 3 seconds (Encoding phase). Then, a 4-second delay was accompanied by a black screen (Maintenance phase). Following the delay, one of the previously presented fractal patterns (target) is shown alongside a novel fractal pattern (foil). The target and foil are displayed along the vertical meridian of the screen, with their relative positions (target above/below foil) randomised across trials (Identification phase). Participants must identify which pattern they just saw (identification performance) by clicking the target pattern and drag it to its proper location on the screen (localisation performance). A demonstration of the OMT is available at: https://octalportal.com/demo-omt/

**Trials**: Participants begin with a practice block of 6 trials (3 one-item trials followed by 3 three-item trials). This is followed by a main test block of 40 trials, consisting of 20 one-item trials and 20 three-item trials. The order of trials within the main block is randomised for each participant. No performance feedback is given during either practice or main test blocks.

**Stimuli:** The stimuli are drawn from a library of 196 fractal images, comprising 49 distinct shapes, each available in 4 colour variations (http://sprott.physics.wisc.edu/fractals.htm). The foil pattern is drawn from the general pool of fractal images but is always one that was not presented as a target in the current trial (i.e., its specific shape and colour combination was not part of the to-be-remembered array for that trial). The on-screen locations for the fractal patterns during encoding are determined pseudo randomly by an offline MATLAB script, with the constraint that fractals are never placed closer than 1.5 times of diameter to one another to avoid visual crowding and to ensure clear zones for localisation error analysis.

##### Outcome metrics

- `OMT_Accuracy` (Identification Accuracy): The proportion of trials where the target fractal is correctly identified. OMT_Acc_Easy means the mean identification accuracy for all easy trials (1 fractal). OMT_Acc_Hard means the mean identification accuracy for all hard trials (3 fractals).
- `OMT_LocError` (Location Error): The Euclidean distance (e.g., in cm or pixels, then scaled) between the centre of the dragged fractal and the centre of its original target location. OMT_LocErr_Easy is the mean across all easy trials. OMT_LocErr_Hard is the mean across all hard trials.
- `OMT_IdeRT` (Identification Time): The reaction time from the presentation of the target-foil pair to the participant’s click identifying the target.
- `OMT_LocRT` (Localisation Time): The reaction time from the initiation of the drag to the placement of the object at its final location.
- `OMT_TargetDetection`: The rate of correctly identifying the target object AND placing it at the correct target location (a composite measure of accuracy).
- `OMT_Misbinding`: The rate of correctly identifying a target but placing it at a location originally occupied by a different non-target item from the encoded array (in 3 fractal trials).
- `OMT_Guessing`: The rate of placing an identified target randomly (“pure guess”), not corresponding to any of the original item locations.

#### Rey-Osterrieth Complex Figure (ROCF)

The ROCF task in OCTAL is a digital adaptation of the traditional pen-and-paper test, designed to assess visuospatial perception, visuoconstructive abilities, and visuospatial episodic memory.^34^ The original ROCF task requires the participant to draw a complex line-drawing freehand, first by replicating an existing figure (copy), and then again from memory (immediate recall). Our digitised version does not require hand drawing. Instead, the figure is split into 13 independent elements, and participants use a drag-and-drop interface to manipulate these elements on an empty digital canvas. The task comprises two phases:

1. Copy Phase: Participants are shown the complete ROCF and are instructed to recreate it by dragging and dropping the predefined elements into their correct positions on the canvas. This phase assesses visuoperceptual organization and visuoconstructive skill.
2. Recall Phase: Immediately following the copy phase, the model figure is removed, and participants are asked to reproduce the figure from memory, again by dragging and dropping the elements. This phase measures visuospatial episodic memory.

A demonstration is available at: https://octalportal.com/demo-rocf/

**Scoring:** An automated, offline scoring algorithm, implemented in MATLAB, is used to evaluate performance, providing a continuous measure of placement precision. This contrasts with the often more subjective, discrete scoring of the traditional version. The scoring logic is as follows:

- Anchor-based referencing: The large, central rectangle of the figure (’a02_large_rectangle’) serves as the primary anchor element. The final (x, y) coordinates of all other placed elements are calculated relative to the centre of this anchor. These relative coordinates are then normalised by the width and height of the anchor element, respectively, to account for variations in the size of the constructed figure. If the primary anchor is not placed, the first element dropped on the canvas is used as a substitute anchor.
- Placement error precision: For each of the 13 elements placed on the canvas, the algorithm calculates the normalised absolute error in both the x and y dimensions. This error is the absolute difference between the element’s final normalised position and its ideal normalised position.
- Decay function: A ‘flat decay’ function is used to convert placement error into a score for each dimension (x and y). An element receives a full score of 1 if its normalised error is within a tolerance of 0.1 (i.e., 10% of the anchor’s dimension). For errors exceeding this tolerance, the score decays, approaching zero for large errors. This method provides credit for approximately correct placements while penalising significant deviations. Alternatively, the scoring function could also be linear function or exponential, where the score for an element decreases sharply and continuously with any deviation from its ideal location. Here we chose to report the data with flat decay to be closer to the pen-and-paper ROCF scoring.
- Total score: The final score for each phase (Copy and Recall) is the sum of the x-scores and y-scores for all placed elements, scaled to a percentage (0–100%). The maximum possible score is reduced for each element that is not moved from the initial ‘pile’, effectively penalising omissions.

##### Outcome metrics

- ROCF_CopyScore, ROCF_RecallScore: A proportional score (0–100%) reflecting the placement accuracy of the figure construction in the copy and recall phases, respectively.
- ROCF_Remember: A memory retention index, calculated as the ratio of ROCF_RecallScore to ROCF_CopyScore (ROCF_RecallScore / ROCF_CopyScore * 100%). This ratio normalises recall for individual differences in baseline visuomotor ability.
- ROCF_CopyDuration: The total time in seconds from the start of the trial to its completion of the copy phase.
- ROCF_RecallDuration: The total time in seconds from the start of the trial to its completion of the recall phase.

#### Object-in-Scene Memory Task (OIS)

The Object-in-Scene (OIS) task is a newly developed memory paradigm, inspired by similar naturalistic object-in-photo tasks,^43^ designed to simultaneously assess both short-term (tested immediately) and long-term (tested after a delay of over one minute) visual associative memory. The task measures multiple components of memory for objects embedded within naturalistic scenes, including identification accuracy, precision of spatial localisation, and semantic memory aspects.

**Procedure:** Participants are presented with a photograph of a natural scene (e.g. a bamboo forest). A specific target object (e.g. a guitar) is shown placed at a random location within this scene. Participants are instructed to remember both the object and its location on the scene. To aid effective encoding, they are required to click on the displayed target object within the scene. Then, the memory is tested in two phases:

- **Immediate recall (short-term memory):** Immediately after encoding an object-scene pair, participants are presented with an array of 20 different objects. They must select the target object they just saw from this array and then drag and place it onto the original scene (now shown without the target object) at its remembered location. To make the object identification component more challenging and to assess semantic interference, the array of 20 objects includes a foil object that is semantically related to the target (e.g., if the target was a specific guitar, the foil might be a different type or colour of guitar).
- **Delayed recall (long-term memory):** After encoding and immediately testing a block of 5 different object-scene pairs, participants undertake a delayed recall phase. In this phase, they are probed on their memory for these 5 pairs, again requiring object identification and localisation. This delayed test typically occurs after an interval of approximately 1-2 minutes.

The task comprises a total of 20 trials (i.e., 20 unique object-scene pairs). These are typically divided into four blocks (e.g., 4 blocks of 5 pairs each for encoding and immediate recall, followed by delayed recall for those blocks). The order of object-scene pairs is randomised for each participant.

A demonstration is available at: https://octalportal.com/demo-ois/

**Stimuli:** The stimuli consist of photographs of everyday scenes and images of various common objects. The object stimuli used in this task were sourced from a standardised set developed by Brady et al. ^44^

**Outcome metrics:** Multiple metrics are extracted for both the immediate (STM) and delayed (LTM) recall phases:

- `OIS_STM_Accuracy` / `OIS_LTM_Accuracy` (Object Identification Accuracy): The proportion of trials in which the participant correctly identifies the original target object from the array. Chance performance for this metric is 5% (1 out of 20).
- `OIS_STM_LocErr` / `OIS_LTM_LocErr` (Location Error): The Euclidean distance, typically measured in centimetres on the calibrated screen, between the centre of the participant’s placed object and the centre of the original target object’s location in the scene.
- Semantic Identification Accuracy (`OIS_STM_SemanticAcc` / `OIS_LTM_SemanticAcc`): The proportion of trials in which the object identified by the participant belongs to the same semantic category as the target object, even if it is not the exact target item (e.g., choosing the foil guitar when the target was a different guitar). Chance performance for this is 10% (assuming two items from the same category are in the array, or as defined by the specific stimulus set construction).
- Identification Reaction Time (`OIS_STM_IdeRT` / `OIS_LTM_IdeRT`): Time taken to identify the object.
- Localisation Reaction Time (`OIS_STM_LocRT` / `OIS_LTM_LocRT`): Time taken to place the object. The OIS task’s use of realistic scenes and objects aims to provide a more ecologically valid assessment of memory. The separation of object identity and spatial location memory, along with STM/LTM distinctions and semantic accuracy, allows for a comprehensive evaluation of visual associative memory.

#### Verbal Memory Wordlist Recall Task (ALF - Accelerated Long-Term Forgetting)

Inspired by the widely used Rey Auditory Verbal Learning Test (RAVLT), the ALF task is designed to assess verbal episodic memory, both reproduction and recognition memory, in immediate recall and delayed recall over 30 minutes delay.

**Procedure**: Participants are presented with a sequence of 12 words. These words are displayed one at a time. Participants are instructed to study the words for a later memory test. Each list of 12 words is constructed such that it contains one word from each of 12 distinct semantic categories (i.e., colour, animals, plants, vegetables, fruits, foods, materials, tools, clothing, parts of a building, weather, occupations). The assignment of words to lists and categories is pseudo-randomised across a large pool of 100 unique word sets. Participants study a given set until a learning criterion of at least 50% correct immediate recall is achieved.

- **Immediate recall**: Immediately after the presentation of the word list (and meeting the encoding criterion), participants engage in two tasks:

- Free recall: They are asked to type or verbally report (if proctored, though OCTAL is remote, so typing is assumed) as many of the 12 words as they can remember, in any order.
- Recognition test: Following free recall, they complete a yes/no recognition test, where they are presented with words and must indicate whether each word was part of the list they just studied.
- **Delayed recall**: Approximately 30 minutes after the immediate test phase, during which other OCTAL tasks are administered, the memory test procedure is repeated. Participants again attempt free recall of the 12-word list and complete another yes/no recognition test for those words.

A demonstration is available at: https://run.pavlovia.org/sijiazhao/demo_alf/

**Stimuli**: The stimuli are lists of 12 common English words, selected to be of simple, high frequency. For scoring of free recall, only exact spellings of the target words are counted as correct (case insensitive). The corpus was selected carefully to make sure that the words are simple and basic (see **Supplementary Table 9** for the full word corpus).

**Outcome metrics**: Performance on the ALF task is quantified by four primary metrics:

- ALF_nWords_immediate: The number of words correctly recalled (exact spelling) during the immediate free recall phase.
- ALF_RecAccuracy_immediate: Accuracy (proportion correct) on the immediate yes/no recognition test.
- ALF_nWords_delayed: The number of words correctly recalled (exact spelling) during the delayed free recall phase (after the 30-minute interval).
- ALF_RecAcc_delayed: Accuracy on the delayed yes/no recognition test.

This task allows for the assessment of initial learning, ensured that the participants, susceptibility to interference over a delay, and the distinction between recall and recognition memory processes. The use of semantically categorised lists also offers the potential for analysing recall strategies (e.g., semantic clustering), though this is not explicitly mentioned as an outcome metric in the provided text.

### Study designs and procedures

The overview of all four studies’ demographics and in-person ACE-III and key OCTAL metrics mean can be found in Table 2.

#### Ethics

Ethical approval for studies involving UK-based participants was granted by the University of Oxford ethics committee (IRAS ID: 248379, Ethics Approval Reference: 18/SC/0448). For Study 1, ethical approval for the recruitment of Chinese participants was granted by the Medical Sciences Interdivisional Research Ethics Committee (MS IDREC) at the University of Oxford (Reference: 671456). All studies were conducted in accordance with the Declaration of Helsinki, and all participants provided written or digital informed consent prior to starting any procedures.

#### Attention check

To ensure data quality from unsupervised remote testing, several task-specific attention checks were embedded for all healthy controls: (1) OIS: Participants were required to drag an object onto the scene immediately after viewing; object identification accuracy <20% (where chance for correct object was 10% and for correct semantic category was 20%) was flagged. No participants failed this check. (2) OMT: An unreasonably short localization time (cut-off at 0.2 seconds) was flagged. Two participants failed this check. (3) DSST: A correct rate <20% (chance for pure guessing ∼11%) or being idle for more than 60 seconds in total was flagged. No participants failed these checks. (3) ROCF Copy: A copy score <80% was flagged. One participant failed this check. (4) Participants who failed only one attention check were retained in the dataset; however, their performance data for the specific task on which the attention check was failed were discarded from analyses involving that task. This strategy balanced the need for data quality with the desire to retain as much valid data as possible.

**Table 2.**
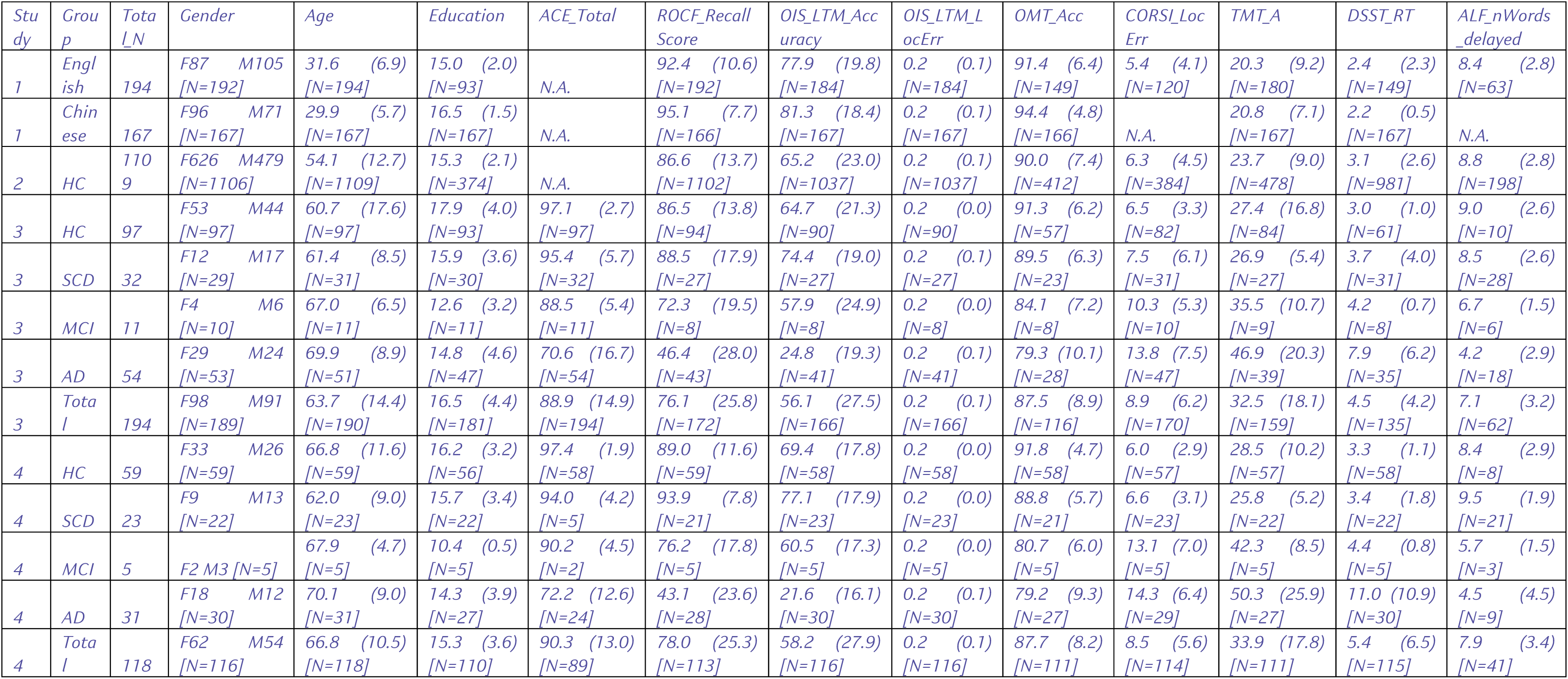
Demographics and ACE, OCTAL performance in four studies. The ACE-III and OCTAL metrics were averaged across assessments for Study 4.

#### Study 1

This study aimed to assess the feasibility and international scalability of the OCTAL platform and to determine whether its cognitive tasks exhibited any inherent bias towards English speakers when administered to young adults from different linguistic backgrounds.

**Participants:** A total of 363 young adults, aged 18–44 years, participated. The sample was divided into two cohorts: (1) English-speaking Cohort: N=194 native English speakers residing in the United Kingdom. Recruitment was conducted via the Prolific online platform. (2) Chinese-speaking Cohort: N=167 native Chinese speakers residing in mainland China. Recruitment was conducted via the Credamo online platform.

Demographic information, including age, gender, and years of education, was collected. For the Chinese cohort, reported education levels were converted to an equivalent years-of-education scale to facilitate cross-cultural comparisons (e.g., ‘Primary school and below’ = ≤6 years, ‘Bachelor’s degree’ = 16 years, ‘Doctoral degree’ = 23 years). The education level options, and year conversions are listed below:

- ‘小学及以下’ (Primary school and below): 6 years or less
- ‘初中’ (Junior high/Middle school): 9 years (6 + 3)
- ‘普高/中专/技校/职高’ (High school/Vocational schools): 12 years (6 + 3 + 3)
- ‘专科’ (Junior college): 15 years (6 + 3 + 3 + 3)
- ‘本科’ (Bachelor’s degree): 16 years (6 + 3 + 3 + 4)
- ‘硕士’ (Master’s degree): 18-19 years (16 + 2-3) :19 (using the higher estimate)
- ‘博士’ (Doctoral degree): 21-23 years (18-19 + 3-4) : 23

**Procedure:** All participants completed the OCTAL battery remotely on their own laptop or desktop computers. The English version of OCTAL was administered to the UK cohort, while a fully translated Chinese version was administered to the China cohort. For a sub-analysis on the effects of screen size, an additional 43 adults aged 45–70 from mainland China were included, expanding this specific analysis cohort to N=210. For this screen size analysis, tablets were disallowed, and the browser automatically captured screen width and height in pixels at the time of testing.

#### Study 2

This study aimed to evaluate the concurrent validity of remote OCTAL assessments by comparing performance with the Addenbrooke’s Cognitive Examination-III (ACE-III), an established in-person cognitive screening tool.

**Participants:** A large cohort of 1,109 UK residents was recruited via Prolific.co. Of these, 905 participants were aged between 45 and 89 years, forming the primary sample for the cognitive aging analyses. Participants self-reported as neurologically healthy, with normal or corrected-to-normal vision, and no colour blindness. To minimize expectancy effects, the study was advertised as a “brain game,” blinding participants to its specific cognitive assessment focus. This sample incorporated 352 healthy adults aged over 50 from a previously published normative study. Recruitment occurred in successive batches to achieve a broad age distribution (overall range 18–85 years), with particular emphasis on obtaining a balanced sample above age 40.

**Procedure**: All assessments were completed remotely and without supervision, using participants’ personal devices. Participants were informed prior to starting that attention checks were embedded within the tasks and that low-effort responses might lead to data rejection.

#### Study 3

This study aimed to evaluate the concurrent validity of remote OCTAL assessments by comparing performance with the Addenbrooke’s Cognitive Examination-III (ACE-III), an established in-person cognitive screening tool.

**Participants:** A total of 194 individuals participated, comprising diverse clinical and control groups: 97 cognitively healthy controls (HC), 54 patients with early Alzheimer’s disease dementia (AD), 11 patients with mild cognitive impairment (MCI), and 32 individuals with subjective cognitive decline (SCD). Patients with AD were recruited from the Cognitive Disorders Clinic at the John Radcliffe Hospital, Oxford, UK. They presented with a progressive, multidomain, predominantly amnestic cognitive impairment, and had undergone MRI and FDG-PET imaging consistent with a clinical diagnosis of AD dementia (e.g., temporo-parietal atrophy and hypometabolism).^45^. All AD, MCI and SCD patients had plasma biomarkers examined, including pTau217, which were in line with their clinical diagnosis. MCI and SCD patients were diagnosed according to Petersen and Jessen’s criteria.^46,47^ HC were recruited from the same clinic as spouses of patients or through open day events. All HCs were aged >50 years, reported no psychiatric or neurological illnesses, were not taking regular psychoactive medications, scored above the cut-off for normality on the ACE-III (≥88/100), and had a normal brain MRI scan as reviewed by two independent senior neurologists (S.T. and M.H.). Participants with colour blindness were excluded at the recruitment stage.

**Procedure:** (1) OCTAL Assessment: Participants completed the OCTAL battery remotely on a personal desktop, laptop, or tablet computer using Chrome or Firefox browsers. A unique, anonymised link containing participant and visit identifiers was emailed to them on the same day as their in-person clinic visit. Participants were encouraged to complete the online tests within one week of the visit. (2) **ACE-III Assessment:** All participants completed the ACE-III in person during their clinic visit, administered by trained personnel as a standard clinical cognitive screening measure. An ACE-III total score <88/100 was considered indicative of cognitive impairment. While HCs scoring below this cut-off were excluded, patients with AD were included based on clinical and radiological criteria, not solely on their ACE-III score. Mild cognitive impairment was defined as ACE-III total score between 87/100 and 83/100.

The order of administration (remote OCTAL vs. in-person ACE-III) was pseudo-balanced across the cohort to control for potential order effects. This design allowed for a direct comparison of a novel remote digital assessment with a traditional, clinician-administered gold-standard screening tool across a spectrum of cognitive abilities.

#### Study 4

This study aimed to quantify the test-retest reliability of the OCTAL battery over a six-month period.

**Participants:** This sample was drawn from the same recruitment pool as Study 3. A cohort of 118 participants was included, representing a similar spectrum of cognitive statuses as in Study 3: 59 cognitively healthy controls (HC), 23 individuals with subjective cognitive decline (SCD), 5 with mild cognitive impairment (MCI), and 31 with Alzheimer’s disease dementia (AD).

**Procedure**: (1) OCTAL Assessments: Participants completed the full OCTAL battery on three separate occasions: at baseline, at 3 months post-baseline, and at 6 months post-baseline. (2) ACE-III Assessments: A subset of 89 participants also completed the paper-based ACE-III at each of these three time points. To minimize practice effects on this comparator measure, alternate forms of the ACE-III (Forms A, B, and C) were used across the assessments. This longitudinal design is essential for establishing the stability of OCTAL metrics over time, a critical property for tools intended for longitudinal monitoring or use in clinical trials. The inclusion of various clinical groups tests the reliability across different levels of cognitive function.

### Statistical analysis

#### General approach

Statistical analyses were performed using MATLAB (version R2025a; MathWorks, Inc.), R (version 12.0; R Core Team), and JASP (version 0.16.4; JASP Team)^48^. Unless otherwise specified, a two-tailed p-value < 0.05 was considered statistically significant. Bonferroni correction for multiple comparisons was applied where appropriate (e.g., for large correlation matrices or multiple group comparisons on various metrics) to control the family-wise error rate. Effect sizes, such as the rank-biserial correlation (r) for Mann-Whitney U tests, were reported to provide information on the magnitude of observed differences between groups.

#### Specific analyses for Study 1

**Group comparisons:** Demographic characteristics and OCTAL performance metrics were compared between the English-speaking and Chinese-speaking cohorts using Mann-Whitney U tests, suitable for two independent groups, particularly when normality of distributions cannot be assumed. Test statistics (U), effect sizes (r), and p-values were reported. Beyond statistical significance, the practical significance of differences in OCTAL metrics between the two language groups was evaluated using a bootstrap permutation procedure with 10,000 iterations. For each metric, the mean performance of the Chinese group was compared against a normative range defined as ±1 standard deviation (SD) from the mean of the English-speaking cohort. A difference was considered practically significant if ≥97.5% of the resampled means from the Chinese group fell below the lower threshold (English Mean −1 SD) or above the upper threshold (English Mean + 1 SD), corresponding to a one-sided p < 0.025. This provides a more robust interpretation of group differences relative to normative variation.

**Screensize correlations:** To investigate the potential influence of display dimensions on OCTAL performance in the remote testing environment, Spearman rank correlations (ρ) were calculated between screen width (and separately, screen height), captured in pixels, and all OCTAL metrics for the expanded Chinese cohort (N=210). P-values for these correlations were Bonferroni-corrected for the number of metrics tested. To formally compare the strength of correlations (e.g., to test if screen width and screen height had significantly different associations with ROCF Copy score), Hittner, May, and Silver’s (2003) modification of Dunn and Clark’s (1969) z-test for dependent overlapping correlations was used, as implemented in the `cocor` package in R.^49–51^

#### Specific analyses for Study 2

**Group comparisons**: Differences in demographic variables between subgroups (e.g., native vs. non-native English speakers) were assessed using Mann-Whitney U tests for continuous variables and chi-squared (χ2) tests for categorical variables.

**Exploratory Factor Analysis (EFA):** To investigate the underlying structure of cognitive performance as measured by OCTAL, an EFA was conducted on 19 key OCTAL metrics from the large healthy cohort (N=1,109). The Kaiser-Meyer-Olkin (KMO) measure of sampling adequacy and Bartlett’s Test of Sphericity (χ2) were used to confirm the suitability of the data for factor analysis. A KMO value of 0.68 indicated acceptable sampling adequacy, and a significant Bartlett’s test supported the factorability of the correlation matrix. Factors were extracted based on eigenvalues >1. An initial analysis used varimax rotation (orthogonal). To allow for potential correlations between cognitive factors, the analysis was repeated using promax rotation (oblique). A χ2 test assessed the sufficiency of the identified number of factors. This EFA helps to understand the main cognitive constructs captured by the OCTAL battery.

**Age trajectories**: To visualise age effects across the lifespan, we applied a year-by-year local smoothing approach. **(1) Normalisation**: Individual participant scores on each OCTAL metric were standardised by converting them to z-scores relative to the mean and standard deviation of a young adult reference group (aged 18–39 years, derived from Study 1 or the younger participants in Study 2). Prior to normalisation, outliers defined as scores >2 SD from the weighted mean at each age point were removed from this specific trajectory analysis to ensure robust trend estimation. The formula used for normalisation was:

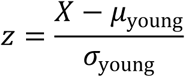

where X represents the OCTAL outcome metric raw value, μ_young_ and σ_young_ represent the mean and standard deviation of the young norm, respectively. For reaction time variables and location error variables, z-scores were sign-reversed so that more negative values consistently indicated poorer performance (i.e., greater decline). **(2) Smoothing**: Year-by-year age trajectories for these normalized scores were generated using a local smoothing technique. Performance at each target age was estimated as a Gaussian-weighted average of participants within a ±5-year window of that age. The weighting function was a Gaussian kernel, defined by a bandwidth (σ) of 2.5 years:

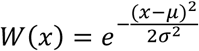

Here, 𝑥𝑥 is the participant’s age and μ is the target age at the centre of the smoothing window. **(3) Linear fits**: To compare the rates of age-related change across different cognitive domains and metrics, straight lines were fitted to the Gaussian-smoothed, age-weighted mean z-scores for each metric (from ages 45 to 75 years) using the `polyfit` function in MATLAB. The slopes of these lines provided an estimate of the average annual change in performance (in z-units per year) and R squared measured the fitness of the linear model (reported in **Supplementary Table 5**).

**Inter-metric correlations**: Spearman rank correlation matrices were computed across all key OCTAL metrics to examine the relationships between different tasks and cognitive domains within the healthy adult sample. Results were presented both with and without Bonferroni correction for multiple comparisons to show the full pattern of relationships and those robust to stringent correction. Correlations between age and individual OCTAL metrics were also calculated using Spearman’s ρ.

**Network module detection:** To investigate the underlying structure of cognitive performance, we constructed a weighted, undirected network from 25 cognitive metrics. The nodes in this network represented each metric, and the edge weights between them were defined by the absolute value of the pairwise Spearman correlation coefficient. We then applied a single-pass Louvain-style modularity maximization algorithm to partition the network into distinct functional communities, or modules.

The Louvain algorithm iteratively optimises network modularity, 𝑄, to find the best partition. Modularity is a scalar value between −0.5 and 1 that measures the density of connections within modules compared to connections between modules. 𝑄 is formally defined as:

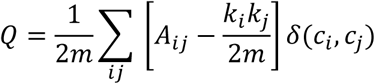

where 𝐴*_ij_* represents the weight of the edge between nodes 𝑖 and 𝑗, 𝑘_𝑖_ and 𝑘_𝑖_ are the sum of weights (strength) of all edges attached to nodes 𝑖 and 𝑗 respectively, 𝑚 is the sum of all edge weights in the network 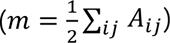, and 𝛿(𝑐_𝑖_, 𝑐_𝑖_) is 1 if nodes 𝑖 and 𝑗 belong to the same community (𝑐_𝑖_ = 𝑐_𝑖_) and 0 otherwise. The algorithm iteratively moves nodes between communities to maximise the gain in 𝑄, and a full pass through all nodes is repeated until no further increase in 𝑄 can be achieved.

To confirm the stability of the detected community structure, we ran the Louvain algorithm for 10,000 iterations. In each iteration, the algorithm’s internal random node ordering allowed for exploration of different local maxima. The modularity 𝑄 for each resultant partition was computed using the formula above. The 6-module solution was robustly obtained in 83.99% of these runs (𝑄 = 0.13 ± 0.00), confirming the high stability of this partition.

Finally, to identify the most influential metric within each community, we calculated the intra-modular strength for every node—defined as the sum of its connection weights to all other nodes within the same module—and designated the node with the highest strength as the central node for that community.

**Language effect analysis**: To explore whether having English as a second language affected OCTAL performance across the lifespan, a non-parametric bootstrap-based resampling method (2,000 iterations) for independent samples was employed. This analysis compared performance between native English speakers and non-native English speakers at each age point, identifying age intervals where significant group differences emerged (defined as p < 0.05, two-tailed, uncorrected for this exploratory analysis).

#### Specific analyses for Study 3

**Age-adjustment of OCTAL metrics**: For comparisons with clinical diagnosis classifications and cognitive impairment defined by ACE-III scores, each participant’s OCTAL metrics were age-adjusted. Each individual’s score was adjusted based on the mean and SD of normative participants from Study 1 and Study 2 within a ±3-year age band around their own age. On average, each individual’s performance was adjusted against 55.8 normative participants.

**Correlations between OCTAL and ACE-III:** Spearman rank correlations were used to assess the association between age-adjusted OCTAL metrics and the ACE-III total score. A Bonferroni-corrected significance level of α=0.0026 was applied for these correlations. To investigate domain-specific relationships, partial Spearman correlations were computed between each OCTAL metric and each of the five ACE-III subscores (Attention, Memory, Fluency, Language, Visuospatial), while statistically controlling for the variance accounted for by the other four ACE-III subscores. This helps to identify unique contributions of OCTAL tasks to specific cognitive domains as defined by the ACE-III.

**Machine learning for feature ranking:** To identify which OCTAL metrics were most predictive of cognitive impairment (defined as an ACE-III total score < 88), a univariate feature ranking approach was used. For the binary classification (impaired vs. unimpaired), chi-square tests (`fscchi2` function in MATLAB) were employed to assess the dependence of each OCTAL metric (feature) on the group classification. The resulting importance scores (-log(p)) were converted to p-values using 𝑃 = 𝑒^−*score*^. For predicting a continuous response variable like the ACE-III total score, F-tests via `fsrftest` in MATLAB was used.

**Receiver Operating Characteristic (ROC) analysis:** ROC analyses were conducted to evaluate the ability of individual OCTAL metrics and combinations of metrics to discriminate between cognitively impaired (ACE-III < 88) and unimpaired (ACE-III ≥ 88) individuals. Area Under the Curve (AUC), sensitivity, specificity, and optimal cut-off thresholds (for both raw scores and age-adjusted z-scores) were determined for key OCTAL metrics and for pre-defined task combinations (e.g., a 20-minute OCTAL subset: ROCF, OIS, TMT; and a 5-minute OCTAL subset: ROCF, DSST).

**Comparison of ROC curves:** DeLong’s test was used to statistically compare the AUCs of different diagnostic models (e.g., to compare the AUC of the 20-minute OCTAL subset against the AUC of the ACE-III total score for distinguishing AD/MCI patients from individuals with SCD).^52^

**Predicting ACE-III scores from OCTAL metrics:** To determine the extent to which remote OCTAL performance could predict scores on a standard clinical assessment, we conducted a series of multiple linear regression analyses using MATLAB (R2025a, MathWorks, Inc.) with the Statistics and Machine Learning Toolbox.

The predictor variables consisted of a subset of nine age-adjusted OCTAL metrics: time to connect two dots (TMT_Connect2), time taken for TMT-A (TMT_A), the TMT B/A ratio (TMT_BdA), ROCF copy and remember scores (ROCF_CopyScore, ROCF_Remember), and object identification accuracy and localisation error for both short- and long-term memory in the OIS task (OIS_STM_Acc, OIS_STM_LocErr, OIS_LTM_Acc, OIS_LTM_LocErr). The outcome variables were the ACE-III total score and its five subscores (Attention, Memory, Visuospatial, Fluency, and Language).

A separate multiple linear regression model (fitlm function in MATLAB) was constructed for each of the six ACE-III outcomes. For each model, participants with incomplete data for any of the included predictor variables or the specific outcome variable were excluded from the analysis on a pairwise basis. The predictive accuracy of each model was assessed by correlating the model-predicted scores with the participants’ actual scores; Pearson’s r was used because it directly quantifies the strength of the linear relationship between the predicted and observed scores, which aligns with the primary objective of a linear regression model to establish the best linear fit to the data. The Mean Absolute Error (MAE) was calculated to quantify the average prediction error.

#### Specific analyses for Study 4

**Test-Retest reliability:** The primary analysis for assessing the six-month test-retest reliability of OCTAL metrics and ACE-III scores was the Intraclass Correlation Coefficient (ICC), originally defined by McGraw & Wong^19^. ICC values were interpreted according to a more recent framework proposed by Koo & Li: values <0.50 were considered poor, 0.50–0.74 as moderate, 0.75–0.90 as good, and >0.90 as excellent reliability.^20^ ICCs, along with their 95% confidence intervals and associated F-statistics, were computed using MATLAB function ICC.^53^ The F-statistics specifically tested the null hypothesis that the true ICC was equal to 0.75 (the threshold for “good” reliability), allowing for a rigorous assessment against this benchmark. This comprehensive approach to reliability assessment is vital for tools intended for repeated measures or longitudinal tracking.

## Acknowledgements

S.Z., S.T., A.S., A.G., M.J.B. and M.H. were funded by the Wellcome Trust (226645/Z/22/Z). S.G.M. was funded by a Medical Research Council (MRC) Clinician Scientist Fellowship (MR/P00878/X) and National Institute of Health and Care Research (NIHR) Oxford Biomedical Research Centre (BRC) and NIHR Oxford Health BRC. Q.-Y. T. was funded by the Guangdong-Hong Kong Universities “1+1+1” Joint Funding Programme. C.G. received a residency scholarship, funded by the Italian Ministry of University and Research (MUR), as part of the Neurology Residency Program at the Univeristà degli Studi di Milano. Thanks to Dr Gabriel Jones for the inspiration of the branding design for OCTAL.

## Authors’ contributions

S.Z., S.To., S.M., and M.H. conceived and designed the study. S.Z. developed and maintained the OCTAL platform. S.Z., S.To., A.S., A.G., C.G., M.J.B., S.M., S.Th., and M.H. were responsible for participant recruitment and data collection. S.Z., S.To., A.S., A.G., and M.J.B. conducted the quality assurance of the remote data. S.Z. curated and analysed the data. S.Z. and M.H. prepared the original manuscript draft. All authors reviewed and edited the paper.

## Data and code availability

The data and code supporting the findings of this study are available to reviewers from the corresponding authors upon request. The code for the OCTAL tasks will be publicly released upon publication of this manuscript, under a free academic licence managed by Oxford University Innovation. Normative data for key OCTAL metrics, stratified by age, will be made publicly available on GitHub (https://github.com/sijiazhao/octal-normative-data), which will be consistently updated as new data are collected. Future releases will aim to include adjustments for gender and education level, subject to data availability. Demonstrations of all OCTAL tasks are currently available at http://octalportal.com.

## Supplementary Materials

### Does speaking English as a second language affect OCTAL performance?

We examined whether speaking English as a second language affects performance on OCTAL, a cognitive battery administered in English.^3^ In our previous study with a smaller sample of participants (Toniolo et al., 2024), we restricted participation to native English speakers. However, in practice—particularly in the UK—many patients maybe fluent in English but speak another language as their mother tongue. Although these individuals may function fluently in English, second-language use can require additional cognitive effort, particularly in verbal tasks. This raises the question: does not having English as a first language impact performance on OCTAL?

In this study, among the 1109 participants, 918 reported English as their first language, while 191 reported a different first language. We hypothesised that the verbal memory task (ALF)—which involves remembering a list of 12 common English words—would be the most susceptible to language-related differences. Surprisingly, no disadvantage was observed for non-native English speakers in either the immediate or delayed recall phases of the ALF task (Figure 7A). In fact, older adults whose first language was not English tended to perform slightly better in both recognition and recall. This may reflect our careful word selection process, which ensured that all ALF items were simple, high-frequency words.

Similarly, we found no evidence of a language effect on performance in other non-verbal tasks, including DSST, ROCF, OMT, and OIS, provided that participants could understand the instructions.

More nuanced effects emerged in the Trail Making Test (TMT). Language background had no impact on the control condition (connecting two dots) or TMT-A (connecting Arabic numerals in sequence), likely because Arabic numerals are universally taught in early education across languages and cultures. However, performance diverged in TMT-B, which requires alternating between numbers and letters (e.g., 1–A–2–B…). From around age 55 onwards, non-native English speakers performed significantly worse than their native English-speaking peers, even when taking TMT-A under control (**see Supplementary Figure 1 below**). Consequently, their TMT-B/A ratio—often used as an index of cognitive flexibility—appeared unreasonably low. This suggests a language-related bias in TMT-B, likely reflecting differences in alphabet familiarity or automaticity rather than true executive dysfunction.

These findings highlight the importance of considering language background when interpreting performance in language-loaded cognitive tasks, particularly when assessing executive function using TMT-B.

**Supplementary Figure 1.**
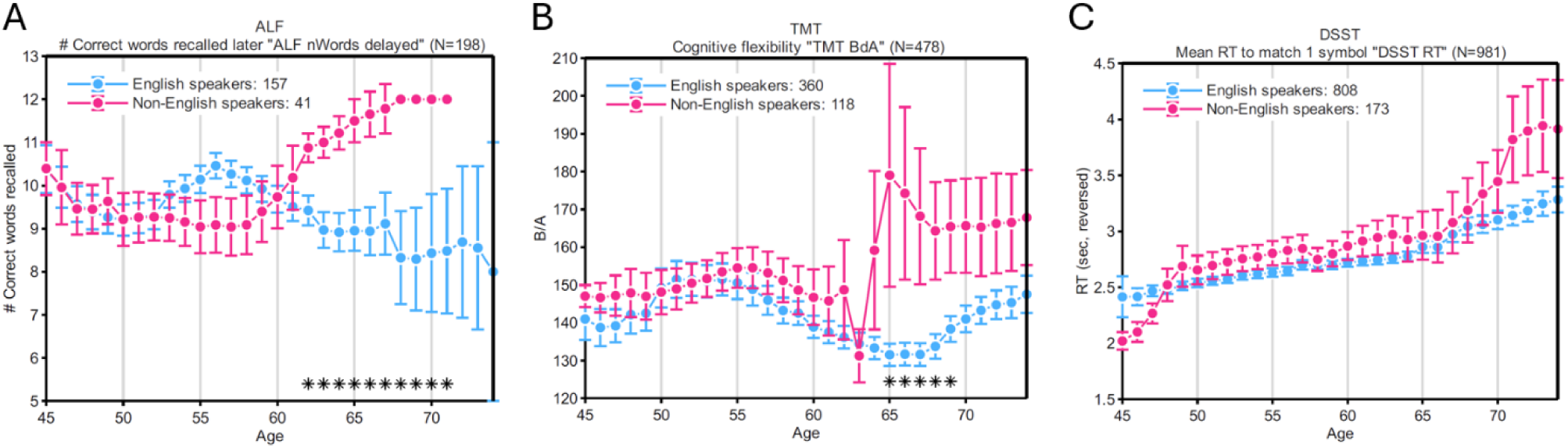
Effect of first language status on OCTAL performance (Study 2, N = 1,109). Scores for native English speakers are plotted in blue, and those for participants who use English as a second language are plotted in bright pink. Asterisks mark age intervals with significant group differences (p < 0.05). Error bars denote ± 1 SEM.

**Supplementary Table 1:**
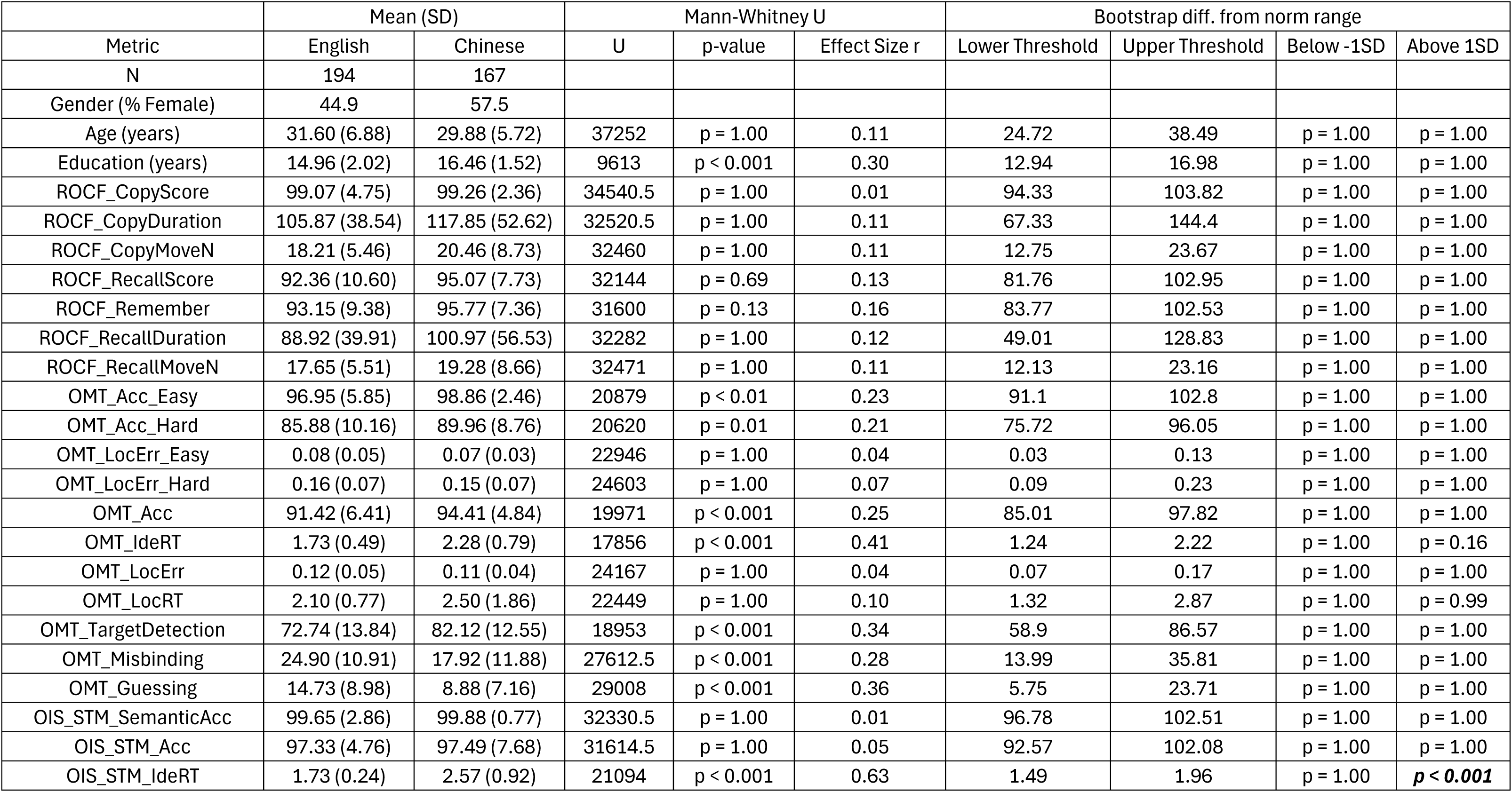
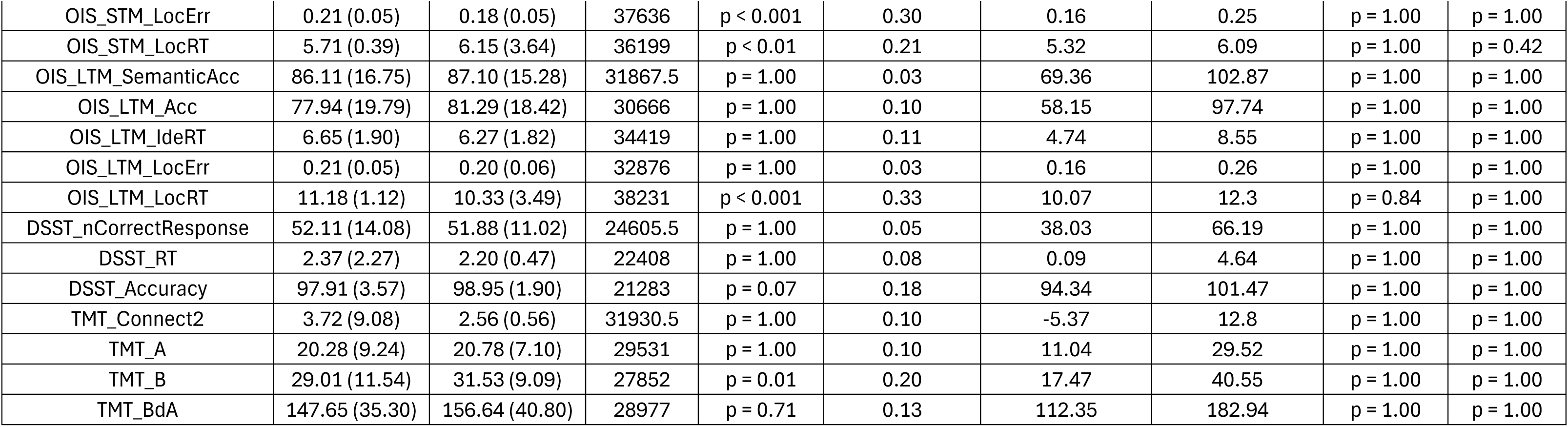
Demographic and cognitive performance comparisons between English- and Chinese-speaking participants (Study 1). Demographic and cognitive performance of English- and Chinese-speaking participants (Study 1). This table extends *Table 1* by reporting Mann–Whitney U statistics, effect sizes (r), and Bonferroni-corrected p values. Practical significance was evaluated with a bootstrap permutation procedure (10,000 iterations). For each metric, lower and upper thresholds were defined as 1 SD below and 1 SD above the mean of the English-speaking cohort, respectively. To determine whether the Chinese group (N = 210) differed meaningfully from these thresholds, N observations were resampled with replacement from the Chinese data, the mean was calculated, and this process was repeated 10,000 times. A difference was deemed significant when ≥97.5 % of resampled means lay below the lower threshold or above the upper threshold (p < 0.025).

| Metric | Mean (SD) |  | Mann-Whitney U |  |  | Bootstrap diff. from norm range |  |  |  |
| --- | --- | --- | --- | --- | --- | --- | --- | --- | --- |
| | English | Chinese | U | p-value | Effect Size $r$ | Lower Threshold | Upper Threshold | Below -1SD | Above 1SD |
| N | 194 | 167 |  |  |  |  |  |  |  |
| Gender (% Female) | 44.9 | 57.5 |  |  |  |  |  |  |  |
| Age (years) | 31.60 (6.88) | 29.88 (5.72) | 37252 | $p = 1.00$ | 0.11 | 24.72 | 38.49 | $p = 1.00$ | $p = 1.00$ |
| Education (years) | 14.96 (2.02) | 16.46 (1.52) | 9613 | $p < 0.001$ | 0.30 | 12.94 | 16.98 | $p = 1.00$ | $p = 1.00$ |
| ROCF_CopyScore | 99.07 (4.75) | 99.26 (2.36) | 34540.5 | $p = 1.00$ | 0.01 | 94.33 | 103.82 | $p = 1.00$ | $p = 1.00$ |
| ROCF_CopyDuration | 105.87 (38.54) | 117.85 (52.62) | 32520.5 | $p = 1.00$ | 0.11 | 67.33 | 144.4 | $p = 1.00$ | $p = 1.00$ |
| ROCF_CopyMoveN | 18.21 (5.46) | 20.46 (8.73) | 32460 | $p = 1.00$ | 0.11 | 12.75 | 23.67 | $p = 1.00$ | $p = 1.00$ |
| ROCF_RecallScore | 92.36 (10.60) | 95.07 (7.73) | 32144 | $p = 0.69$ | 0.13 | 81.76 | 102.95 | $p = 1.00$ | $p = 1.00$ |
| ROCF_Remember | 93.15 (9.38) | 95.77 (7.36) | 31600 | $p = 0.13$ | 0.16 | 83.77 | 102.53 | $p = 1.00$ | $p = 1.00$ |
| ROCF_RecallDuration | 88.92 (39.91) | 100.97 (56.53) | 32282 | $p = 1.00$ | 0.12 | 49.01 | 128.83 | $p = 1.00$ | $p = 1.00$ |
| ROCF_RecallMoveN | 17.65 (5.51) | 19.28 (8.66) | 32471 | $p = 1.00$ | 0.11 | 12.13 | 23.16 | $p = 1.00$ | $p = 1.00$ |
| OMT_Acc_Easy | 96.95 (5.85) | 98.86 (2.46) | 20879 | $p < 0.01$ | 0.23 | 91.1 | 102.8 | $p = 1.00$ | $p = 1.00$ |
| OMT_Acc_Hard | 85.88 (10.16) | 89.96 (8.76) | 20620 | $p = 0.01$ | 0.21 | 75.72 | 96.05 | $p = 1.00$ | $p = 1.00$ |
| OMT_LocErr_Easy | 0.08 (0.05) | 0.07 (0.03) | 22946 | $p = 1.00$ | 0.04 | 0.03 | 0.13 | $p = 1.00$ | $p = 1.00$ |
| OMT_LocErr_Hard | 0.16 (0.07) | 0.15 (0.07) | 24603 | $p = 1.00$ | 0.07 | 0.09 | 0.23 | $p = 1.00$ | $p = 1.00$ |
| OMT_Acc | 91.42 (6.41) | 94.41 (4.84) | 19971 | $p < 0.001$ | 0.25 | 85.01 | 97.82 | $p = 1.00$ | $p = 1.00$ |
| OMT_IdeRT | 1.73 (0.49) | 2.28 (0.79) | 17856 | $p < 0.001$ | 0.41 | 1.24 | 2.22 | $p = 1.00$ | $p = 0.16$ |
| OMT_LocErr | 0.12 (0.05) | 0.11 (0.04) | 24167 | $p = 1.00$ | 0.04 | 0.07 | 0.17 | $p = 1.00$ | $p = 1.00$ |
| OMT_LocRT | 2.10 (0.77) | 2.50 (1.86) | 22449 | $p = 1.00$ | 0.10 | 1.32 | 2.87 | $p = 1.00$ | $p = 0.99$ |
| OMT_TargetDetection | 72.74 (13.84) | 82.12 (12.55) | 18953 | $p < 0.001$ | 0.34 | 58.9 | 86.57 | $p = 1.00$ | $p = 1.00$ |
| OMT_Misbinding | 24.90 (10.91) | 17.92 (11.88) | 27612.5 | $p < 0.001$ | 0.28 | 13.99 | 35.81 | $p = 1.00$ | $p = 1.00$ |
| OMT_Guessing | 14.73 (8.98) | 8.88 (7.16) | 29008 | $p < 0.001$ | 0.36 | 5.75 | 23.71 | $p = 1.00$ | $p = 1.00$ |
| OIS_STM_SemanticAcc | 99.65 (2.86) | 99.88 (0.77) | 32330.5 | $p = 1.00$ | 0.01 | 96.78 | 102.51 | $p = 1.00$ | $p = 1.00$ |
| OIS_STM_Acc | 97.33 (4.76) | 97.49 (7.68) | 31614.5 | $p = 1.00$ | 0.05 | 92.57 | 102.08 | $p = 1.00$ | $p = 1.00$ |
| OIS_STM_IdeRT | 1.73 (0.24) | 2.57 (0.92) | 21094 | $p < 0.001$ | 0.63 | 1.49 | 1.96 | $p = 1.00$ | **$p < 0.001$** |
| OIS_STM_LocErr | 0.21 (0.05) | 0.18 (0.05) | 37636 | p < 0.001 | 0.30 | 0.16 | 0.25 | p = 1.00 | p = 1.00 |
| OIS_STM_LocRT | 5.71 (0.39) | 6.15 (3.64) | 36199 | p < 0.01 | 0.21 | 5.32 | 6.09 | p = 1.00 | p = 0.42 |
| OIS_LTM_SemanticAcc | 86.11 (16.75) | 87.10 (15.28) | 31867.5 | p = 1.00 | 0.03 | 69.36 | 102.87 | p = 1.00 | p = 1.00 |
| OIS_LTM_Acc | 77.94 (19.79) | 81.29 (18.42) | 30666 | p = 1.00 | 0.10 | 58.15 | 97.74 | p = 1.00 | p = 1.00 |
| OIS_LTM_IdeRT | 6.65 (1.90) | 6.27 (1.82) | 34419 | p = 1.00 | 0.11 | 4.74 | 8.55 | p = 1.00 | p = 1.00 |
| OIS_LTM_LocErr | 0.21 (0.05) | 0.20 (0.06) | 32876 | p = 1.00 | 0.03 | 0.16 | 0.26 | p = 1.00 | p = 1.00 |
| OIS_LTM_LocRT | 11.18 (1.12) | 10.33 (3.49) | 38231 | p < 0.001 | 0.33 | 10.07 | 12.3 | p = 0.84 | p = 1.00 |
| DSST_nCorrectResponse | 52.11 (14.08) | 51.88 (11.02) | 24605.5 | p = 1.00 | 0.05 | 38.03 | 66.19 | p = 1.00 | p = 1.00 |
| DSST_RT | 2.37 (2.27) | 2.20 (0.47) | 22408 | p = 1.00 | 0.08 | 0.09 | 4.64 | p = 1.00 | p = 1.00 |
| DSST_Accuracy | 97.91 (3.57) | 98.95 (1.90) | 21283 | p = 0.07 | 0.18 | 94.34 | 101.47 | p = 1.00 | p = 1.00 |
| TMT_Connect2 | 3.72 (9.08) | 2.56 (0.56) | 31930.5 | p = 1.00 | 0.10 | -5.37 | 12.8 | p = 1.00 | p = 1.00 |
| TMT_A | 20.28 (9.24) | 20.78 (7.10) | 29531 | p = 1.00 | 0.10 | 11.04 | 29.52 | p = 1.00 | p = 1.00 |
| TMT_B | 29.01 (11.54) | 31.53 (9.09) | 27852 | p = 0.01 | 0.20 | 17.47 | 40.55 | p = 1.00 | p = 1.00 |
| TMT_BdA | 147.65 (35.30) | 156.64 (40.80) | 28977 | p = 0.71 | 0.13 | 112.35 | 182.94 | p = 1.00 | p = 1.00 |

**Supplementary Figure 2:**
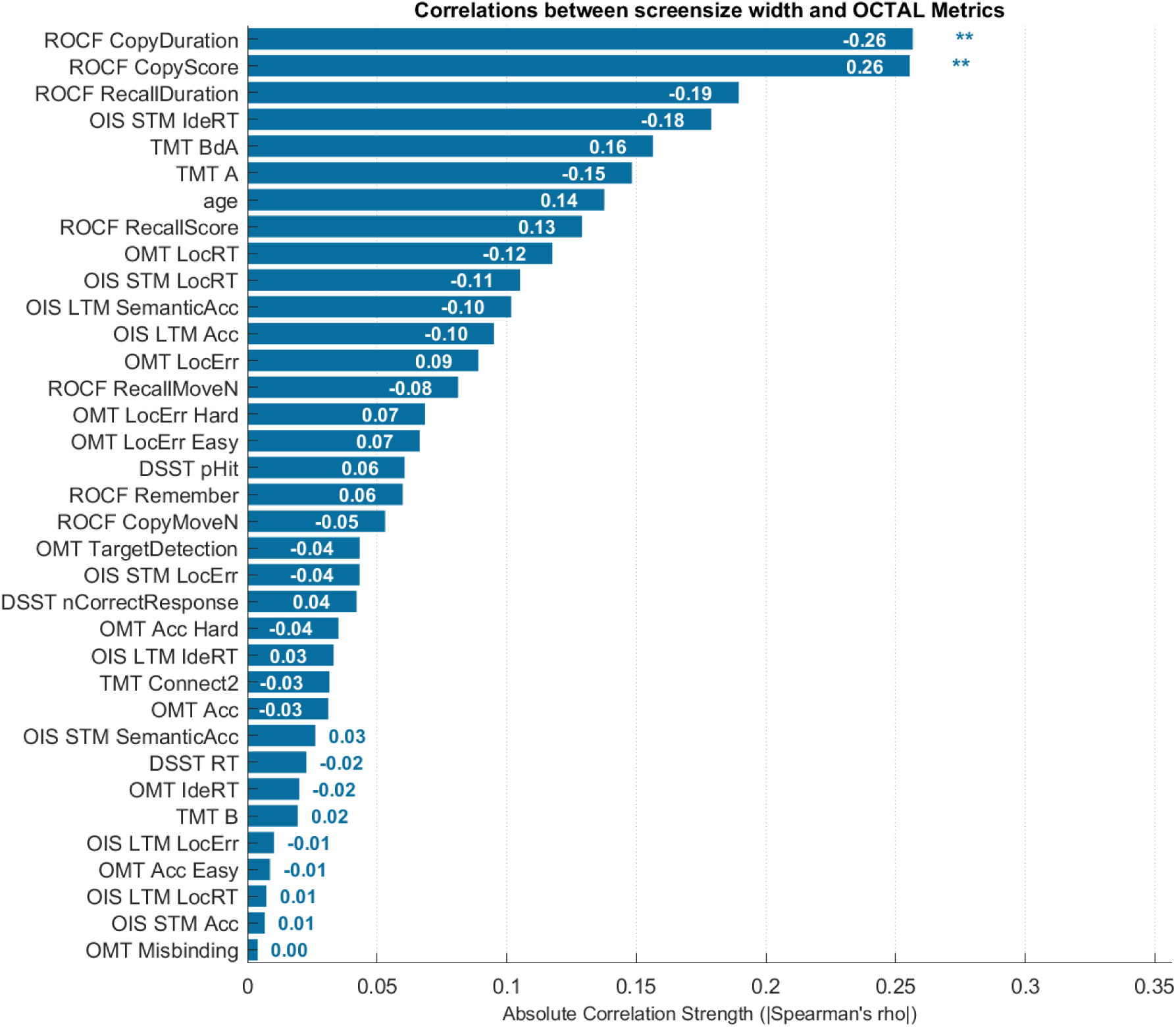
Rank-ordered Spearman correlations between screen width and OCTAL task metrics. P values were Bonferroni-corrected for 35 comparisons (34 OCTAL metrics + age); ** denotes p < 0.01. The white numerals at the ends of bars gives the spearman rho values. All correlations were computed from a sample of N=210 in Study 1. The width of screen was captured and recorded automatically by the Credamo platform. The details of p values are available in **Supplementary Table 2**.

**Supplementary Table 2:** Spearman correlations between screen width and OCTAL metrics.

| *Metric* | *rho* | *pValue* | *Significance* |
| --- | --- | --- | --- |
| *ROCF_CopyDuration* | -0.26 | 0.01 | ** |
| *ROCF_CopyScore* | 0.26 | 0.01 | ** |
| *ROCF_RecallDuration* | -0.19 | 0.21 |  |
| *OIS_STM_IdeRT* | -0.18 | 0.33 |  |
| *TMT_BdA* | 0.16 | 0.83 |  |
| *TMT_A* | -0.15 | 1.00 |  |
| *age* | 0.14 | 1.00 |  |
| *ROCF_RecallScore* | 0.13 | 1.00 |  |
| *OMT_LocRT* | -0.12 | 1.00 |  |
| *OIS_STM_LocRT* | -0.11 | 1.00 |  |
| *OIS_LTM_SemanticAcc* | -0.10 | 1.00 |  |
| *OIS_LTM_Acc* | -0.10 | 1.00 |  |
| *OMT_LocErr* | 0.09 | 1.00 |  |
| *ROCF_RecallMoveN* | -0.08 | 1.00 |  |
| *OMT_LocErr_Hard* | 0.07 | 1.00 |  |
| *OMT_LocErr_Easy* | 0.07 | 1.00 |  |
| *DSST_pHit* | 0.06 | 1.00 |  |
| *ROCF_Remember* | 0.06 | 1.00 |  |
| *ROCF_CopyMoveN* | -0.05 | 1.00 |  |
| *OMT_TargetDetection* | -0.04 | 1.00 |  |
| *OIS_STM_LocErr* | -0.04 | 1.00 |  |
| *DSST_nCorrectResponse* | 0.04 | 1.00 |  |
| *OMT_Acc_Hard* | -0.04 | 1.00 |  |
| *OIS_LTM_IdeRT* | 0.03 | 1.00 |  |
| *TMT_Connect2* | -0.03 | 1.00 |  |
| *OMT_Acc* | -0.03 | 1.00 |  |
| *OIS_STM_SemanticAcc* | 0.03 | 1.00 |  |
| *DSST_RT* | -0.02 | 1.00 |  |
| *OMT_IdeRT* | -0.02 | 1.00 |  |
| *TMT_B* | 0.02 | 1.00 |  |
| *OIS_LTM_LocErr* | -0.01 | 1.00 |  |
| *OMT_Acc_Easy* | -0.01 | 1.00 |  |
| *OIS_LTM_LocRT* | *0.01* | *1.00* |  |
| *OIS_STM_Acc* | *0.01* | *1.00* |  |
| *OMT_Misbinding* | *0.00* | *1.00* |  |

**Supplementary Figure 3:**
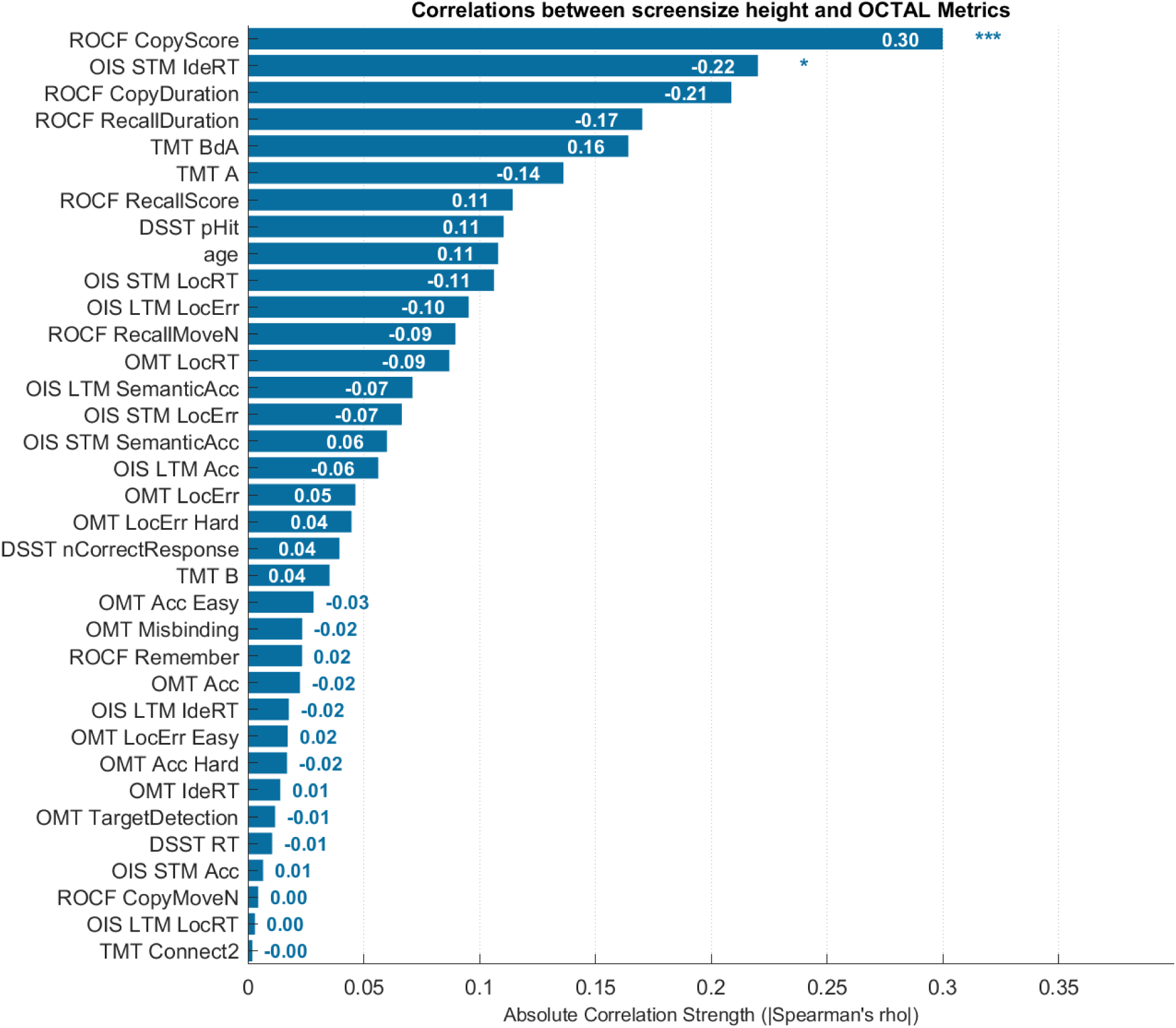
Rank-ordered Spearman correlations between screen height and OCTAL task metrics. P values were Bonferroni-corrected for 35 comparisons (34 OCTAL metrics + age);* denotes p < 0.05, *** denotes p < 0.001. The white numerals at the ends of bars gives the spearman rho values. All correlations were computed from a sample of N=210 in Study 1. The width of screen was captured and recorded automatically by the Credamo platform. The details of p values are available in **Supplementary Table 3**.

**Supplementary Table 3:** Spearman correlations between screen height and OCTAL metrics.

| *Metric* | *rho* | *pValue* | *Significance* |
| --- | --- | --- | --- |
| *ROCF_CopyScore* | 0.30 | 0.0004 | *** |
| *OIS_STM_IdeRT* | -0.22 | 0.05 | * |
| *ROCF_CopyDuration* | -0.21 | 0.08 |  |
| *ROCF_RecallDuration* | -0.17 | 0.48 |  |
| *TMT_BdA* | 0.16 | 0.61 |  |
| *TMT_A* | -0.14 | 1.00 |  |
| *ROCF_RecallScore* | 0.11 | 1.00 |  |
| *DSST_pHit* | 0.11 | 1.00 |  |
| *age* | 0.11 | 1.00 |  |
| *OIS_STM_LocRT* | -0.11 | 1.00 |  |
| *OIS_LTM_LocErr* | -0.10 | 1.00 |  |
| *ROCF_RecallMoveN* | -0.09 | 1.00 |  |
| *OMT_LocRT* | -0.09 | 1.00 |  |
| *OIS_LTM_SemanticAcc* | -0.07 | 1.00 |  |
| *OIS_STM_LocErr* | -0.07 | 1.00 |  |
| *OIS_STM_SemanticAcc* | 0.06 | 1.00 |  |
| *OIS_LTM_Acc* | -0.06 | 1.00 |  |
| *OMT_LocErr* | 0.05 | 1.00 |  |
| *OMT_LocErr_Hard* | 0.04 | 1.00 |  |
| *DSST_nCorrectResponse* | 0.04 | 1.00 |  |
| *TMT_B* | 0.04 | 1.00 |  |
| *OMT_Acc_Easy* | -0.03 | 1.00 |  |
| *OMT_Misbinding* | -0.02 | 1.00 |  |
| *ROCF_Remember* | 0.02 | 1.00 |  |
| *OMT_Acc* | -0.02 | 1.00 |  |
| *OIS_LTM_IdeRT* | -0.02 | 1.00 |  |
| *OMT_LocErr_Easy* | 0.02 | 1.00 |  |
| *OMT_Acc_Hard* | -0.02 | 1.00 |  |
| *OMT_IdeRT* | 0.01 | 1.00 |  |
| *OMT_TargetDetection* | -0.01 | 1.00 |  |
| *DSST_RT* | -0.01 | 1.00 |  |
| *OIS_STM_Acc* | 0.01 | 1.00 |  |
| *ROCF_CopyMoveN* | *0.00* | *1.00* |  |
| *OIS_LTM_LocRT* | *0.00* | *1.00* |  |
| *TMT_Connect2* | *0.00* | *1.00* |  |

**Supplementary Figure 4:**
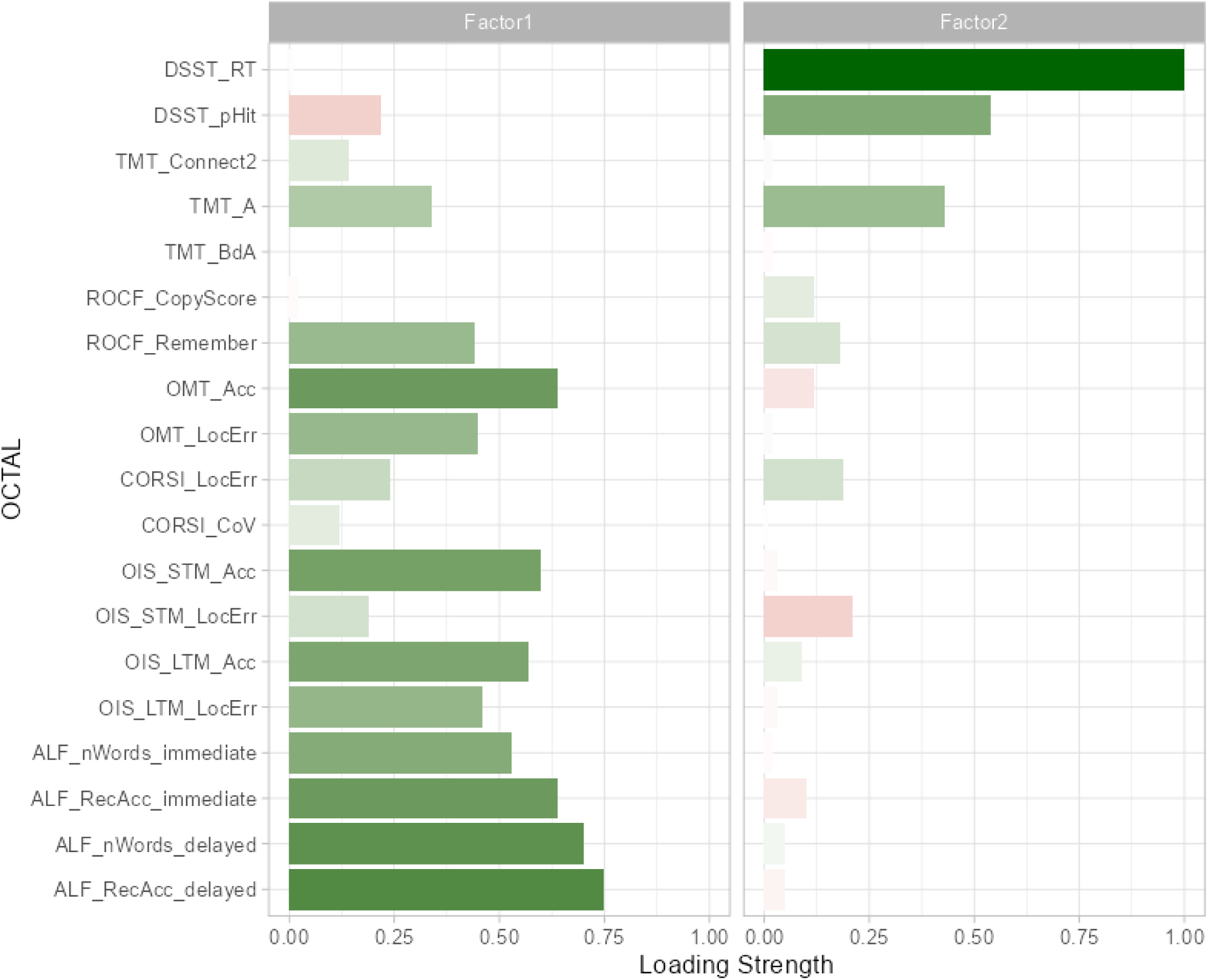
Two-factor structure derived from an exploratory factor analysis of 19 OCTAL metrics obtained remotely from 1,109 healthy participants across the adult lifespan (Study 2). Item labels show task abbreviation and metric; their order is arbitrary. Bar colour indicates the sign of the factor loading (pink = negative, green = positive). Factor 1 (memory) is dominated by high loadings from ROCF Recall, OMT, OIS, and ALF measures of both accuracy and localisation, whereas Factor 2 (executive function) loads chiefly on DSST and TMT-A indices.

**Supplementary Table 4:** Spearman correlations between age and OCTAL metrics in Study 2.

| *Metric* | *N* | *rho* | *p* | *Significance* |
| --- | --- | --- | --- | --- |
| *TMT_A* | 478 | 0.46 | $p < 0.001$ | *** |
| *TMT_B* | 478 | 0.40 | $p < 0.001$ | *** |
| *DSST_nCorrectResponse* | 987 | -0.39 | $p < 0.001$ | *** |
| *OMT_IdeRT* | 420 | 0.39 | $p < 0.001$ | *** |
| *TMT_Connect2* | 478 | 0.33 | $p < 0.001$ | *** |
| *OIS_STM_IdeRT* | 1039 | 0.31 | $p < 0.001$ | *** |
| *OMT_LocRT* | 420 | 0.28 | $p < 0.001$ | *** |
| *ROCF_RecallScore* | 1102 | -0.27 | $p < 0.001$ | *** |
| *OIS_LTM_Acc* | 1037 | -0.26 | $p < 0.001$ | *** |
| *OIS_LTM_SemanticAcc* | 1037 | -0.25 | $p < 0.001$ | *** |
| *CORSI_LocErr* | 384 | 0.24 | $p < 0.001$ | *** |
| *ROCF_CopyScore* | 1101 | -0.23 | $p < 0.001$ | *** |
| *ROCF_Remember* | 1101 | -0.19 | $p < 0.001$ | *** |
| *ROCF_Remember* | 1101 | -0.19 | $p < 0.001$ | *** |
| *ALF_RecAcc_immediate* | 198 | 0.19 | $p = 0.009$ | |
| *OIS_LTM_IdeRT* | 1039 | 0.18 | $p < 0.001$ | *** |
| *OMT_Acc* | 412 | -0.16 | $p = 0.002$ | |
| *OMT_Misbinding* | 412 | 0.15 | $p = 0.002$ | |
| *OMT_Acc_Hard* | 412 | -0.15 | $p = 0.002$ | |
| *DSST_pHit* | 981 | -0.15 | $p < 0.001$ | *** |
| *OIS_STM_LocErr* | 1039 | -0.14 | $p < 0.001$ | *** |
| *OIS_STM_Acc* | 1039 | -0.14 | $p < 0.001$ | *** |
| *OMT_TargetDetection* | 420 | -0.11 | $p = 0.026$ | |
| *OMT_LocErr_Easy* | 412 | -0.11 | $p = 0.030$ | |
| *ALF_nWords_delayed* | 198 | 0.10 | $p = 0.141$ | |
| *ROCF_RecallDuration* | 1102 | 0.10 | $p < 0.001$ | * |
| *ALF_nWords_immediate* | 198 | 0.09 | $p = 0.218$ | |
| *OIS_STM_LocRT* | 1039 | 0.09 | $p = 0.005$ | |
| *OMT_Acc_Easy* | 412 | -0.08 | $p = 0.095$ | |
| *ROCF_CopyDuration* | 1101 | 0.08 | $p = 0.011$ | |
| *OMT_LocErr_Hard* | 412 | 0.07 | $p = 0.162$ | |
| *OIS_LTM_LocRT* | 1039 | 0.06 | $p = 0.062$ | |
| *ALF_RecAcc_delayed* | *198* | *-0.05* | *p = 0.521* |  |
| *CORSI_CoV* | *384* | *-0.04* | *p = 0.444* |  |
| *TMT_BdA* | *478* | *-0.01* | *p = 0.770* |  |
| *OIS_STM_SemanticAcc* | *1039* | *-0.01* | *p = 0.823* |  |
| *ROCF_RecallMoveN* | *1102* | *0.01* | *p = 0.864* |  |
| *OMT_LocErr* | *412* | *0.00* | *p = 0.928* |  |
| *ROCF_CopyMoveN* | *1101* | *0.00* | *p = 0.964* |  |
| *OIS_LTM_LocErr* | *1037* | *0.00* | *p = 0.974* |  |

**Supplementary Figure 5:**
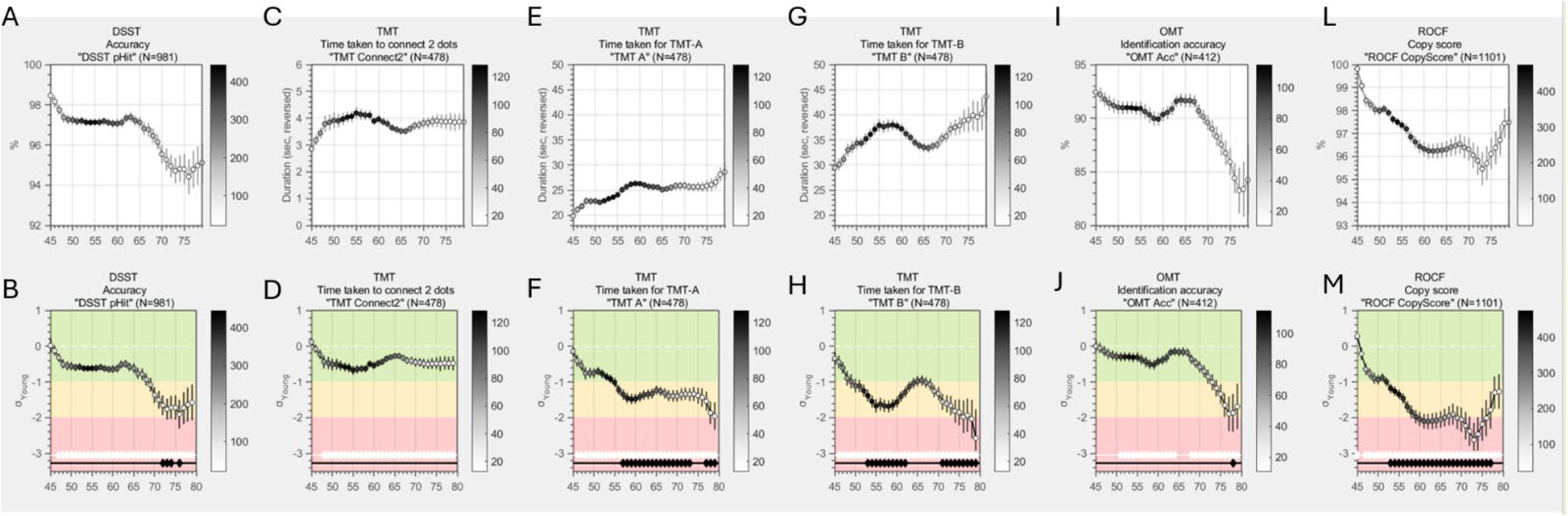
Age trajectories of OCTAL metrics in a healthy population (Study 2, N = 1,109). This is additional to Figure 2 in the main manuscript. The top panel displays raw scores for the DSST (A), OIS Short-Term (C) and Long-Term (E) recall, and ROCF memory (G) across ages 45–80 years. Curves are smoothed with a Gaussian kernel (bandwidth = 2.5 years). Marker colour encodes the number of observations contributing to each point, as shown by the colour bar on the right. The bottom panel presents the same metrics normalised to the young-adult reference group (18–39 years). The scale demarcates the normative range (±1 SD, green zone), moderate deviation (1–2 SD, orange zone), and marked impairment (≥2 SD, red zone). Reaction-time variables—such as DSST mean RT in panel A—are sign-reversed so that increasingly negative z-scores indicate greater executive decline. White diamonds along the horizontal white guideline mark age groups that differ significantly from the young-adult mean, whereas black diamonds atop black stems indicate significant deviation from –1 SD (i.e., the boundary between the green and orange zones).

**Supplementary Table 5:** Slopes of age-effect on OCTAL metrics. This is supplementary to Figure 3 in the main text.

| *Metric* | *Slope (z/year)* | *R squared* |
| --- | --- | --- |
| *ROCF_CopyScore* | -0.07034 | 0.832969 |
| *DSST_RT* | -0.0726 | 0.948665 |
| *TMT_Connect2* | -0.00924 | 0.332046 |
| *TMT_A* | -0.03229 | 0.666922 |
| *TMT_B* | -0.02339 | 0.296664 |
| *TMT_BdA* | 0.010007 | 0.129192 |
| *OMT_Acc* | -0.02427 | 0.493306 |
| *OIS_STM_Acc* | -0.01469 | 0.474455 |
| *OIS_LTM_Acc* | -0.03358 | 0.683793 |
| *OIS_Remember* | -0.03115 | 0.659559 |
| *ROCF_Remember* | -0.03503 | 0.811077 |
| *ALF_nWords_immediate* | -0.00894 | 0.146788 |
| *ALF_nWords_delayed* | -0.01987 | 0.70015 |
| *ALF_Remember* | -0.01745 | 0.632817 |

**Supplementary Figure 6.**
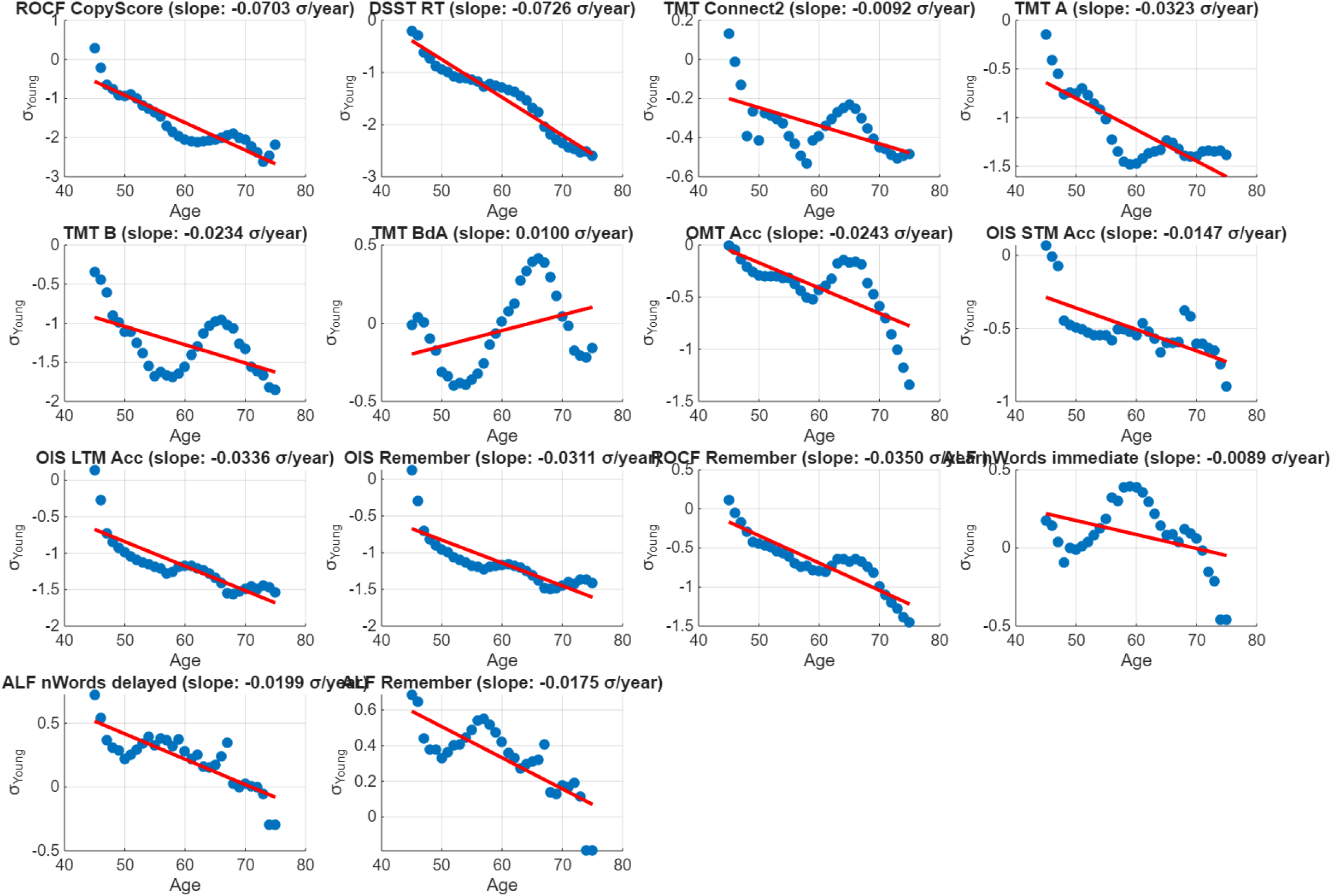
Scatter plots depicting the linear relation between age and each OCTAL metric. Individual participant scores (N = 1,109) are shown together with least-squares regression lines, providing the numerical slopes that underpin the ranked trajectories in Figure 3. The blue dots showing the kernel-smoothed normalised metrics against age. The red line shows the regression line, with the slope value shown in the title.

**Supplementary Figure 7:**
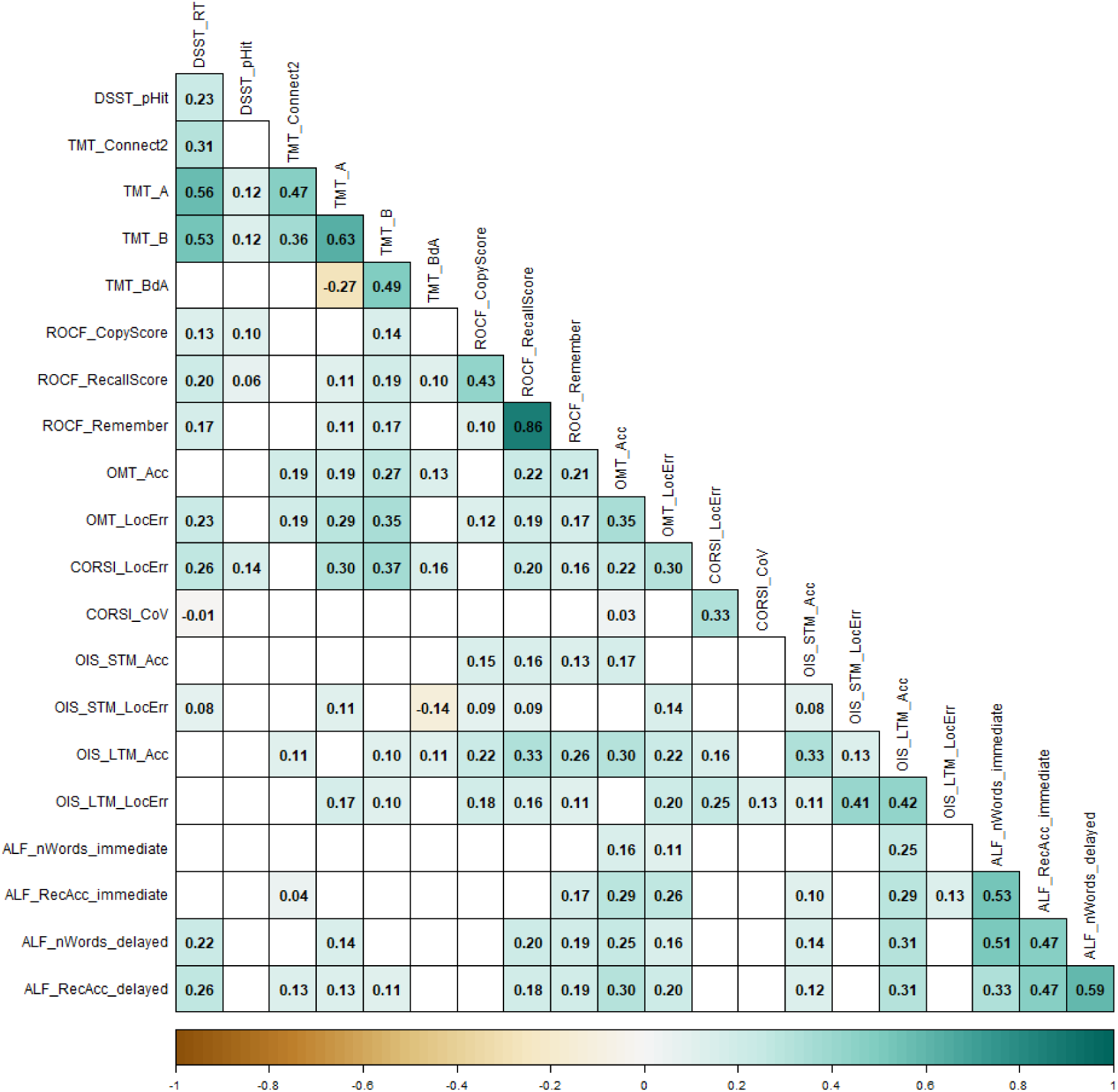
Metric-by-metric correlations among OCTAL indices in healthy adults (Study 2), without multiple comparison correction. The matrix displays Spearman coefficients (ρ) for all pairwise comparisons; only significant correlations (p < 0.05) are shown. All metrics are directionally aligned so that lower scores indicate greater cognitive impairment. Square colour, mapped to the accompanying colour bar, encodes the magnitude and sign of ρ, and each cell is labelled with its exact value.

**Supplementary Table 6:** Correlation table between ACE-III subscores. All values are Spearman’s rho and all significant (p < 0.001 after Bonferroni correction).

|  | ACE-III Attention | ACE-III Memory | ACE-III Visuospatial | ACE-III Fluency | ACE-III Language |
| --- | --- | --- | --- | --- | --- |
| ACE-III Attention | 1.00 | 0.65 | 0.64 | 0.57 | 0.44 |
| ACE-III Memory | 0.65 | 1.00 | 0.61 | 0.69 | 0.55 |
| ACE-III Visuospatial | 0.64 | 0.61 | 1.00 | 0.56 | 0.52 |
| ACE-III Fluency | 0.57 | 0.69 | 0.56 | 1.00 | 0.52 |
| ACE-III Language | 0.44 | 0.55 | 0.52 | 0.52 | 1.00 |

**Supplementary Figure 8:**
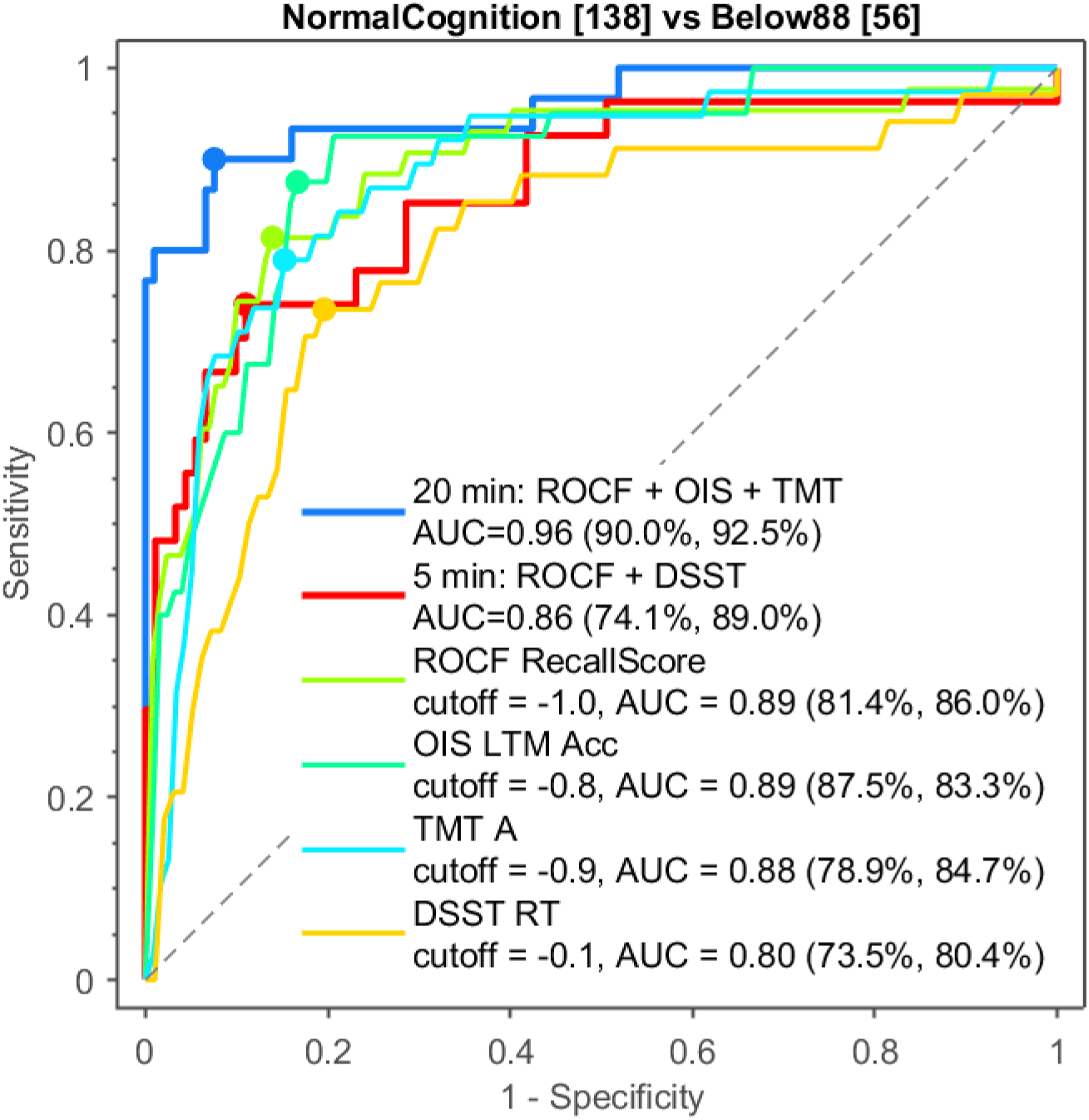
ROC for zscore.

**Supplementary Table 7:** Linear regression model of ROCF + OIS + TMT.

| Row | Estimate | SE | tStat | pValue |
| --- | --- | --- | --- | --- |
| Intercept | 3.518326 | 7.629387 | 0.461155 | 0.644688 |
| ROCF_CopyScore | -0.06508 | 0.071634 | -0.90844 | 0.363644 |
| ROCF_CopyMoveN | -0.06786 | 0.055741 | -1.21746 | 0.223429 |
| ROCF_RecallScore | -0.04205 | 0.022713 | -1.85118 | 0.064143 |
| ROCF_RecallMoveN | -0.04852 | 0.041748 | -1.16212 | 0.245185 |
| OIS_STM_Acc | 0.001786 | 0.033698 | 0.053008 | 0.957726 |
| OIS_LTM_Acc | -0.06131 | 0.02553 | -2.40144 | 0.016331 |
| TMT_A | -0.02462 | 0.023569 | -1.04472 | 0.296151 |
| TMT_BdA | -0.0209 | 0.009078 | -2.30191 | 0.02134 |

**Supplementary Table 8:** Intraclass correlation coefficients (ICCs) for all OCTAL and ACE-III metrics. Two-sided 95 % confidence intervals are given in square brackets. The accompanying F-statistics test the null hypothesis that the ICC equals 0.75—the threshold for “good” reliability under the interpretative framework of Koo and Li^20^ —at an α level of 0.05.

| *Metric* | *ICC* | *F<sub>stat</sub>* |
| --- | --- | --- |
| *DSST Accuracy* | 1.00 [1.00 1.00] | $F(111,222) = 162.59, p < 0.001$ |
| *OMT Acc* | 0.98 [0.98 0.99] | $F(89,178) = 19.01, p < 0.001$ |
| *TMT BdA* | 0.98 [0.97 0.99] | $F(108,216) = 14.53, p < 0.001$ |
| *ACE-III Total* | 0.98 [0.97 0.98] | $F(120,240) = 11.86, p < 0.001$ |
| *TMT B* | 0.97 [0.96 0.98] | $F(108,216) = 10.73, p < 0.001$ |
| *OMT TargetDetection* | 0.96 [0.95 0.97] | $F(89,178) = 7.70, p < 0.001$ |
| *ALF nWords delayed* | 0.96 [0.94 0.98] | $F(52,104) = 7.51, p < 0.001$ |
| *ROCF CopyScore* | 0.96 [0.95 0.97] | $F(148,296) = 7.16, p < 0.001$ |
| *OMT Guessing* | 0.95 [0.94 0.97] | $F(89,178) = 6.36, p < 0.001$ |
| *OMT Misbinding* | 0.95 [0.93 0.97] | $F(89,178) = 6.15, p < 0.001$ |
| *OIS LTM LocErr* | 0.95 [0.93 0.96] | $F(132,264) = 5.89, p < 0.001$ |
| *ACE-III Memory* | 0.95 [0.93 0.96] | $F(120,240) = 5.42, p < 0.001$ |
| *ALF nWords immediate* | 0.92 [0.88 0.95] | $F(52,104) = 3.79, p < 0.001$ |
| *ACE-III Fluency* | 0.92 [0.89 0.94] | $F(120,240) = 3.48, p < 0.001$ |
| *DSST RT* | 0.92 [0.89 0.94] | $F(110,220) = 3.46, p < 0.001$ |
| *ACE-III Attention* | 0.91 [0.89 0.94] | $F(120,240) = 3.30, p < 0.001$ |
| *ROCF RecallScore* | 0.90 [0.88 0.93] | $F(147,294) = 2.92, p < 0.001$ |
| *OIS STM LocErr* | 0.90 [0.87 0.92] | $F(131,262) = 2.77, p < 0.001$ |
| *TMT A* | 0.89 [0.85 0.92] | $F(108,216) = 2.55, p < 0.001$ |
| *ACE-III Visuospatial* | 0.89 [0.85 0.92] | $F(120,240) = 2.48, p < 0.001$ |
| *CORSI LocErr* | 0.88 [0.85 0.91] | $F(142,284) = 2.41, p < 0.001$ |
| *ROCF Remember* | 0.87 [0.84 0.90] | $F(146,292) = 2.19, p < 0.001$ |
| *TMT Connect2* | 0.87 [0.83 0.91] | $F(108,216) = 2.17, p < 0.001$ |
| *OIS LTM Acc* | 0.86 [0.82 0.89] | $F(133,266) = 1.92, p < 0.001$ |
| *CORSI CoV* | 0.85 [0.80 0.88] | $F(142,284) = 1.74, p < 0.001$ |
| *OIS STM Acc* | 0.82 [0.77 0.86] | $F(131,262) = 1.45, p = 0.006$ |
| *ACE-III Language* | 0.80 [0.75 0.85] | $F(120,240) = 1.32, p = 0.035$ |
| *OMT LocErr* | 0.79 [0.72 0.85] | $F(89,178) = 1.21, p = 0.139$ |

**Supplementary Table 9:** The full word corpus used in Verbal Memory Wordlist Recall Task (ALF). The first row lists all 12 semantic categories. In the main task, one word would be randomly selected from each category.

| ***colour*** | ***animals*** | ***plants*** | ***vegetables*** | ***fruits*** | ***foods*** | ***materials*** | ***tools*** | ***clothing*** | ***parts of a building*** | ***weather*** | ***occupations*** |
| --- | --- | --- | --- | --- | --- | --- | --- | --- | --- | --- | --- |
| *red* | *tiger* | *tulip* | *spinach* | *apple* | *garlic* | *gold* | *spoon* | *shoes* | *wall* | *rain* | *engineer* |
| *orange* | *lion* | *rose* | *bean* | *lemon* | *oil* | *silver* | *fork* | *blouse* | *floor* | *snow* | *doctor* |
| *yellow* | *horse* | *lily* | *potato* | *banana* | *sugar* | *copper* | *pot* | *pants* | *roof* | *storm* | *lawyer* |
| *green* | *dog* | *daisy* | *carrot* | *grape* | *salt* | *wood* | *pan* | *skirt* | *ceiling* | *cloud* | *pilot* |
| *blue* | *cat* | *tree* | *lettuce* | *peach* | *tea* | *glass* | *pen* | *shirt* | *window* | *sun* | *teacher* |
| *purple* | *cow* | *grass* | *corn* | *orange* | *coffee* | *stone* | *knife* | *socks* | *door* | *fog* | *farmer* |
| *brown* | *turtle* | *ivy* | *pepper* | *berry* | *wine* | *plastic* | *cup* | *scarf* | *basement* | *ice* | *driver* |
| *white* | *frog* | *bamboo* | *pea* | *melon* | *bread* | *paper* | *bowl* | *hat* | *loft* | *rainbow* | *dentist* |
| *black* | *fish* | *mushroom* | *tomato* | *pear* | *milk* | *steel* | *plate* | *tie* | *garden* | *wind* | *nurse* |

## Footnotes

1 The version reported in this manuscript was for laptops/computers or tablets. We now have optimised task for a smartphone version called miniOCTAL, which features the same task design, but is shorter and more suitable for portrait layout. The validation of the smartphone version is still ongoing and will be reported in a separate work.

3 The Chinese-language OCTAL battery excluded the Verbal Memory Word-List Recall task because text-entry methods are not standardised across age groups in China: younger adults typically use pinyin keyboards, whereas many older adults rely on stroke-based input (e.g., Wubi), handwriting interfaces, or have limited typing skills, preventing equitable administration.

3 OCTAL is also available in Simplified Chinese, normative data is collecting in the mainland of China, but this is not included in this manuscript.

